# Contextual factors and implementation strategies for a multi-level community-based sodium reduction intervention in Chicago’s South Side: A qualitative study

**DOI:** 10.1101/2024.10.21.24315888

**Authors:** Olutobi A. Sanuade, Allison J. Carroll, Ricky Watson, Jiancheng Ye, Jennie Hill, Jonathan Chipman, Fernando Wilson, Andy J. King, Abel Kho, Guilherme Del Fiol, Paris Davis, Justin D. Smith

## Abstract

**Background:** Excessive sodium intake exacerbates rates of hypertension. African American adults have higher rates of hypertension in part due to a higher-sodium diet. The multi-level Communication for Behavioral Impact for Sodium Reduction (COMBI-SR) community-based intervention effectively reduces sodium intake in international settings, but it has not yet been implemented and tested in the U.S. This study explored the contextual factors (barriers/facilitators) and implementation strategies for COMBI-SR in Chicago’s South Side neighborhood–an area with high rates of hypertension.

**Methods:** Data were gathered between May and November 2023, through qualitative interviews and focus groups with 40 local participants, including community members, faith-based leaders, and healthcare professionals. The Consolidated Framework for Implementation Research (CFIR) 2.0 guided the development of semi-structured interview guides. Thematic analysis was performed with coding based on CFIR 2.0 constructs (contextual factors) and the Expert Recommendations for Implementing Change (ERIC) compilation of implementation strategies.

**Results:** Key barriers included a lack of awareness of sodium content in foods, socioeconomic disparities limiting access to healthy options, and cultural dietary traditions. Facilitators included strong community partnerships, engaged faith-based organizations, and openness to integrating technology, such as a mobile app, to help monitor and reduce sodium intake. Specific strategies to support sodium reduction involved simplifying public health messages, offering low sodium cooking demonstrations, promoting healthier food options through community outreach, and providing personalized education on reading nutrition labels and managing sodium intake.

**Conclusion:** Successful implementation of COMBI-SR in Chicago’s South Side requires addressing financial, educational, and cultural barriers while leveraging trusted community structures to promote sustainable sodium reduction. These findings will guide future efforts to implement COMBI-SR in the U.S. and improve cardiovascular health.

## Background

Almost 1 in 3 American adults has hypertension.^1^ About three-quarter of U.S. adults with hypertension do not have their BP under control, increasing their risk for heart disease and stroke.^1^ Hypertension contributes to 685,875 deaths annually in the United States (U.S.), and disproportionately affects African American (AA) adults.^2^ The 30-year gap in life expectancy between Chicago’s South Side predominantly AA neighborhoods, for example, and the northern, predominately white neighborhoods is partly attributable to hypertension rates^3,4^—78.7% in the South Side compared to 21.7% in the northern neighborhoods.^5^

Excess sodium intake is associated with high BP and increased risk of heart disease and stroke.^6,7^ Research shows that the largest number of diet-related deaths globally (1.89 million deaths) is associated with excess sodium intake.^8^ The American Heart Association (AHA) recommends sodium intake of <2300 mg/day to reduce the risk of cardiovascular disease, yet, the average American consumes about 3,400 mg daily, nearly 50% above the recommended level.^9^ Reducing sodium intake to AHA-recommended levels could prevent thousands of deaths annually, significantly reduce the prevalence of high BP, improve cardiovascular health, and save $18 billion/year in healthcare costs.^10^

Communication for Behavioral Impact for Sodium Reduction (COMBI-SR) is an evidence-based multi-level intervention designed to empower people to improve their diets and increase demand for lower-sodium food products.^11^ COMBI-SR includes both individual- and community-level approaches. At the individual level, tools like the SaltSwitch smartphone application (app) help users to change their behaviors and decrease personal sodium intake.^12^ At the community level, mass media communication campaigns, community outreach, and partnerships with local healthcare and government organizations drive shifts in the culture surrounding sodium knowledge and intake. Previous trials showed that COMBI-SR effectively reduced sodium consumption, lowered BP, and improved knowledge and behaviors regarding sodium consumption.^11,13,14^ These studies took place in Vietnam and Australia; thus, the intervention likely needs adaptation to local contexts to be effective and sustainable in the U.S. This study explores the contextual factors (i.e., barriers and facilitators) and strategies for implementing the COMBI-SR program in Chicago’s South Side neighborhood. The findings will guide the development of context-specific adaptations of COMBI-SR for this community.

## Methods

### Study design and settings

A qualitative study was conducted to understand the barriers and facilitators to reducing sodium intake (i.e., salt use during cooking, salt use at the table, selection of low salt processed foods) and implementation strategies for addressing these determinants in Chicago’s South Side neighborhood. This study was reported according to the Consolidated Criteria for Reporting Qualitative Research (COREQ). Institutional Review Board of Northwestern University gave ethical approval for this work (STU00212622).

### Intervention

The COMBI-SR program is an evidence-based multi-level intervention that promotes sodium reduction through both individual- and community-level approaches. ^11,13,14^ The individual-level intervention is the SaltSwitch smartphone app, which is designed to help users to access sodium content in processed foods and find lower-sodium alternatives. Community-level interventions include mass media campaigns, outreach efforts, and partnerships with local healthcare and government organizations to drive broader engagement.^11,13,14^

### Study participants

Participants were purposively sampled from Chicago’s South Side community for their membership in one of three groups: potential intervention recipients, research ministry ambassadors (RMAs), and healthcare professionals. Recruitment was conducted in partnership with Pastors for Patient-Centered Outcomes Research (Pastors4PCOR), a Patient-Centered Outcomes Research Institute (PCORI)-funded community health outreach initiative that engages a diverse range of faith communities in Chicago’s South Side and south suburbs to foster partnerships between community members and researchers to address health disparities. The three groups of participants described below provided diverse perspectives on the context and strategies for sodium reduction in Chicago’s South Side community. Participants received incentives for their time: $150 gift cards for potential intervention recipients and RMAs, $250 gift cards for healthcare professionals.

#### Potential intervention recipients

We recruited community members living with hypertension and/or caring for someone with hypertension from Faith-Based Organizations (FBOs) on the South Side of Chicago. FBOs, often trusted within African American communities, provide structured health promotion activities including fairs, screenings, or education, often referred to as health ministries.^15^ These participants shared their perspectives on sodium intake, daily recommendations, public health messaging, use of nutrition technology platforms, health impacts, and community resources.

#### Research ministry ambassadors (RMAs)

RMAs are leaders in Chicago’s South Side faith-based communities who underwent research and community engagement training with Pastors4PCOR. They facilitate engagement across diverse faith communities and offer researcher workshops on strategies and methods for engaging both academic and medical research communities. These participants shared their perspectives on sodium intake, daily recommendations, public health messaging, use of nutrition technology platforms, and health impacts of sodium. They also provided insights on their willingness (as an RMA) to promote salt reduction, past advocacy efforts, potential champions for COMBI-SR, and community resources needed for successful implementation.

#### Healthcare professionals

We recruited healthcare professionals (physicians, nurse practitioners, community healthcare workers, pharmacists, clinic administrators, and physician assistants/primary care providers) from AllianceChicago–a health center-controlled and practice-based research network committed to advancing health equity and fostering innovative collaboration. The network connects over 50 organizations, serving more than 3.5 million patients across Illinois and 19 other states; these participants were affiliated with Federally Qualified Health Centers (FQHCs) with clinics located in Chicago’s South Side. These participants provided their perspectives on conducting brief lifestyle interventions, such as advice, encouragement, or communication, which research suggests may help individuals monitor their sodium intake, use food/nutrition labels, and choose lower-sodium foods.^16^

### Interviews and focus groups

Data were collected though in-depth interviews and focus groups between May and November 2023 by four authors with qualitative research experience (OAS, JY, RW, and RJ). Fifteen in-depth interviews (approximately 30 minutes each) were conducted with potential intervention recipients (n=8), RMAs (n=5) and healthcare professionals (n=2). Four focus groups (approximately 90 minutes each) were conducted, including 1 group of potential intervention recipients (n=9) and 3 groups of healthcare professionals (n=10).

Semi-structured interview guides were developed using the updated Consolidated Framework for Implementation Research (CFIR 2.0)^17^ and modified based on discussions with Pastors4PCOR leadership. The interview guides were tailored to ask about specific COMBI-SR components that were relevant to each group. The potential intervention recipient guide focused on their general attitudes towards sodium consumption and awareness of daily sodium intake recommendations. The RMA guide focused on their general attitudes towards sodium consumption, and awareness of daily sodium intake recommendations, effectiveness of public health messaging on sodium, familiarity with and use of technology platforms for nutrition information, health impacts of excessive sodium consumption, willingness to promote salt reduction, past advocacy efforts, potential champions for COMBI-SR, and the community resources needed for successful implementation. The healthcare professional guide focused on barriers to and facilitators of sodium reduction, strategies for reducing sodium intake, and the role of healthcare professionals in addressing excess sodium consumption in their patient population. Details of the interview guides are provided in **Supplemental File 1**. All interviews and focus groups were conducted in English and recorded via the Zoom videoconferencing platform.

### Data analysis

All interviews and focus groups were transcribed verbatim, redacted for confidentiality, and uploaded to Dedoose v9.0.17 software for coding and analyses.^18^ Transcripts were analyzed thematically using a deductive approach, and guided by the CFIR 2.0^17^ and the Expert Recommendation for Implementing Change (ERIC) compilation.^19^ Coding was performed by two authors (OAS and AJC), who collaboratively developed the codebook by first double-coding 3 transcripts (1 RMA interview, 1 FBO focus group, and 1 health professional focus group) to agree on key themes and optimize inter-coder reliability. The remaining transcripts were divided between the two coders for independent coding and were cross-checked and discussed. Participant characteristics and survey responses (healthcare professionals only) were summarized as frequency and percentage.

## Results

### Participant Characteristics

**Table 1** provides the characteristics of the study participants. Of the participants interviewed, 65% were female, and 75% identified as Black/African American. All potential intervention recipients and RMAs were Black/African American (100%). All potential intervention recipients and RMAs were Black/African American (100%). Healthcare professionals were more racially diverse, with 66.7% identifying as White. The mean age of healthcare professionals was 45.3 years (SD = 7.9), with 91.7% holding a college or postgraduate degree, and most reported an annual household income between $100,000 and $199,999 (72.7%).

**Table 1.** Participants characteristics.

|  | **Total**<br>**(N=40)** | **Potential**<br>**Intervention**<br>**Recipients (n=19)** | **Research Ministry**<br>**Ambassadors**<br>**(n=9)** | **Healthcare**<br>**Professionals**<br>**(n=12)** |
| --- | --- | --- | --- | --- |
|  | **n (%)** | **n (%)** | **n (%)** | **n (%) or M (SD)** |
| **Sex** |  |  |  |  |
| Male | 14 (35.0) | 8 (42.1) | 4 (44.4) | 11 (91.7) |
| Female | 26 (65.0) | 11 (57.9) | 5 (55.6) | 1 (8.3) |
| **Race** |  |  |  |  |
| Asian | 1 (2.5) | 0 (0.0) | 0 (0.0) | 1 (8.3) |
| Black/African American | 30 (75.0) | 19 (100.0) | 9 (100.0) | 2 (16.7) |
| White | 8 (20.0) | 0 (0.0) | 0 (0.0) | 8 (66.7) |
| Other | 1 (2.5) | 0 (0.0) | 0 (0.0) | 1 (8.3) |
| Age, years | - | - | - | 45.3 (7.9) |
| **Education** |  |  |  |  |
| <College | - |  |  | 1 (8.3) |
| College/postgrad | - |  |  | 11 (91.7) |
| **Annual household income** |  |  |  |  |
| <\$100,000 | - | - | - | 2 (18.2) |
| \$100,000-\$199,999 | - | - | - | 8 (72.7) |
| \$200,000+ | - | - | - | 1 (9.1) |

Potential Intervention Recipients and Research Ministry Ambassadors were not asked to report age, education and annual household income.

### Overall Context of Sodium Consumption

Both potential intervention recipients and RMAs expressed confusion about the difference between sodium and salt, with some associating salt with unhealthy habits and sodium with fitness or nutritional benefits. Nonetheless, all participants generally viewed excess salt or sodium as harmful, associating it with negative health conditions such as high BP, headaches, and body swelling. Some participants (RMAs, potential intervention recipients) were unaware that sodium is a nutrient, and many associated it exclusively with processed foods, junk food, and fried items, and viewed it as entirely negative. Many admitted to being unfamiliar with sodium consumption guidelines and did not know the recommended daily intake. However, a few of the potential intervention recipients recognized that certain types of salt could cure headaches, and others highlighted its role as a flavor/taste enhancer. Additionally, a biblical reference to salt as a form of punishment was noted by some participants, while some respondents who self-identified as fitness enthusiasts saw sodium as essential for energy and workout performance.

### Determinants of Implementing COMBI-SR

The following results are presented per the five CFIR 2.0 domains: Innovation, Outer setting, Inner setting, Individuals, and Implementation Process. Details of the codes applied are presented in **Supplemental File 2**. Details of the constructs and corresponding themes and quotes are presented in **Supplemental File 3**.

#### Innovation

Innovation is the evidence-based “thing” being implemented —in this study, the innovation is the SaltSwitch app (individual level) and COMBI-SR messaging (community level). While some found the dietary messages of COMBI-SR to be clear and effective, others felt that overly complex information could create resistance (*Innovation Complexity*). Regarding SaltSwitch, participants noted the need for a user-friendly app with straightforward instructions and language to make the app accessible; concerns were raised about difficulties with reading food labels and confusion about sodium content (*Innovation Complexity*).

Participants expressed an inclination to use and promote SaltSwitch, particularly if it was offered for free. The appeal of “free” was noted to transcend demographic boundaries (*Innovation Cost*). Participants likewise stated that the cost of lower-sodium alternatives recommended by SaltSwitch should be modest to ensure accessibility.

Participants responded positively to COMBI-SR’s public health message examples *(Innovation Design)*, appreciating vibrant design and clear depictions (e.g., ‘5 grams’ as one teaspoon). They suggested including more information about the link between sodium intake and heart disease to better understand its relevance to their health. The SaltSwitch app was also well-received for offering easy access to nutritional information and providing healthier alternatives, though participants recommended improving usability in selecting the best healthier alternatives. (*Innovation Design*).

Many of the potential intervention recipients and RMAs raised concerns about the sufficiency of available research about the role of salt intake on physical health, suggesting that more research may be necessary to fully understand this association, particularly on conditions like high BP (*Innovation Evidence Base*). Participants desired clearer explanations about how salt intake recommendations are developed, especially given individual differences in salt sensitivity. Additionally, participants sought greater transparency in research methodologies and inclusivity to ensure sodium intake guidelines are accurate and relevant for diverse groups.

> *So like I said, for me, in my opinion, I still think that, as it relates to biology as it relates to food, we still need a hundred more, a hundred to 400 more years to really speak accurately about like hat salt is really doing in someone’s body, instead of just going with the hypothesis like, Oh, this person eats too much fried chicken with salt, and that’s the reason why they developed this (FBO 4)*

Finally, participants preferred tools that enhance health consciousness and simplify nutrition decision-making. They reported using lifestyle and fitness apps and social media to track nutrition, exercise, weight loss and to detect health issues. Compared to similar apps, participants expressed interest in adopting SaltSwitch if it is free or more affordable, provides real-time lower-sodium alternatives, and simplifies label interpretation for informed nutritional choices *(Innovation Relative Advantage*). Notably, participants emphasized that SaltSwitch must offer lower sodium alternatives that maintain good taste to be enticing for users.

#### Outer Setting

Outer setting is the context in which the Inner Setting exists —here, Outer Setting includes community/neighborhood factors (outside the faith-based community) driving implementation of sodium reduction interventions in Chicago’s South Side.

Participants demonstrated varied awareness of sodium intake in the community, which was compounded by generational dietary practices (*Local Attitudes)*. Participants further highlighted the cost of healthier foods as a significant barrier to a healthy lifestyle *(Local Conditions)*. They noted that while fast-food restaurants offer healthier choices today, the primary issue remains the higher cost of healthier foods compared to cheaper, less healthy options. Participants described how lack of knowledge about healthy options, inferior nutritional quality of food bank donations, and pervasive presence of sodium in daily products (e.g., toothpaste) are barriers to reducing sodium intake.

Participants recommended leveraging a variety of community resources to promote healthier eating habits, such as: integrating healthy food options into shelters, distributing fresh produce through food pantries, partnering with local businesses, and utilizing educational institutions, faith-based organizations, healthcare settings, and community kitchens to spread nutrition knowledge and support healthier dietary practices *(Partnerships & Connections)*.

Participants identified policies and community incentives to promote healthy eating and address food insecurity, such as enhancing the Special Supplemental Nutrition Program for Women, Infants, and Children (WIC) program, increasing government commitment to population health, and ensuring sustainable policies beyond temporary grants (*Policies & Laws)*. Moreover, funding food and nutrition initiatives as essential healthcare, and advocating for the same level of support as pharmaceutical companies to improve public health, are key *(Financing)*. Lastly, participants highlighted the barrier to healthier food options posed when businesses prioritize their financial gain over the nutritional content of their products *(External Pressure)*.

#### Inner Setting

Inner Setting is the setting in which the innovation is implemented (e.g., hospital, school, city)—here, the Inner Setting includes the FBOs and community organizations with whom we will partner to implement COMBI-SR in Chicago’s South Side.

Potential intervention recipients and RMAs admitted to not fully understanding how much salt or sodium they consume, often overlooking the sodium content in items like salad dressings and processed snacks (*Access to Knowledge and Information)*. They cited insufficient education on the consequences of sodium consumption, particularly the extent of its presence in food and associated health implications (e.g., hypertension, cardiovascular disease, gout), especially in communities with less access to comprehensive nutritional education.

Lack of financial backing was cited as barriers to dietary program sustainability, nutrition class access, and effective health education and nutrition management broadly *(Available Resources)*. Other noted resource limitations included store closures with healthier options (e.g., citing that Whole Foods location on the South Side had closed. Participants acknowledged opportunities in the rise of health-conscious dining options and the expanding availability of organic, farm-raised, and pesticide-free products.

All participants emphasized that community networks play a crucial role in managing health by promoting issues (e.g., high BP, headaches) via regular medical follow-ups, community health metrics, and support networks (*Communications*). Key communication channels for promoting healthy diet practices included online support groups and family discussions. RMAs and healthcare professionals noted the absence of targeted salt reduction programs and the difficulty of delivering comprehensive dietary education during brief clinical visits as significant challenges (*Compatibility*). They also discussed struggles with maintaining long-term engagement in health initiatives, citing a sugar reduction program that failed to sustain participant interest despite initial enthusiasm.

Potential intervention recipients and RMAs commented on the multifaceted role of salt in faith-based communities, highlighting its dual symbolism in religious contexts and cultural significance (*Culture*). For example, the positive connotation of the phrase, “salt of the earth,” is counter to the negative connotation of the story of Lot’s wife: The story of Lot’s wife comes from the Bible, where she and her family were instructed by angels to flee the city of Sodom without looking back as it was being destroyed. However, Lot’s wife disobeyed, turned to look back, and was turned into a pillar of salt as punishment. All participants described deep-rooted cultural traditions surrounding the use of salt in communal meals and noted resistance to change, particularly when it comes to altering traditional, generational recipes.

RMAs highlighted the shift among Pastors4PCOR and individual FBOs from a focus on sugar to embracing broader health initiatives, noting that sodium intake would be aligned with the overarching commitment, purpose, and goal of their community (*Mission Alignment*). For example, RMAs described health literacy campaigns and events like “Man Church,” which addressed prostate cancer and mental health.

Food deserts, safety concerns, and transportation concerns were structural barriers identified as limiting access to fresh food and inadvertently directing people toward unhealthy options (*Structural Characteristics*). Compared to other community health initiatives (e.g., depression, substance use), participants viewed dietary concerns and programs such as COMBI-SR as less critical (*Relative priority)*. They also critiqued food labels, noting that calories are displayed prominently, connoting greater importance, while sodium, sugar, and other nutritional details are in smaller print.

#### Individuals

The Individuals domain refers to the roles and characteristics (Need, Capability, Opportunity, Motivation) of individuals involved in implementation—here, we focused on Innovation Deliverers (e.g., RMAs, healthcare professionals) and Innovation Recipients (e.g., patients, community members). Note, only characteristics of Capability and Motivation were coded for Innovation Deliverers.

Potential intervention recipients were deeply aware of the negative health consequences of excessive salt consumption, such as hypertension and heart disease, often via family anecdotes as well as direct health experiences (e.g., observing the effects of salt intake, such as body swelling or health improvements after dietary changes) (*Need: Innovation Recipients*). In contrast, factors that hindered efforts to reduce salt intake included the convenience and widespread availability of fast food, absence of healthier options, and deeply rooted cultural food practices that are difficult to change.

The narratives of potential intervention recipients and RMAs revealed varied levels of awareness, intentionality, and confidence regarding their salt consumption (*Capability: Innovation Recipients*). Some of them expressed a lack of knowledge about sodium content in foods, challenges in identifying low-sodium alternatives, and difficulty in and doubts about changing long-standing dietary habits. Other participants demonstrated capacity to reduce their salt intake, such as using alternative seasonings (e.g., garlic or onion powder) and being mindful of portion sizes.

Healthcare professionals and RMAs largely expressed confidence in their ability to educate and positively influence people’s dietary behaviors (*Capability: Innovation Deliverers*). However, some mentioned challenges that patients face such as a lack of proficiency and confidence in understanding and managing sodium intake. Participants noted that their efforts would be facilitated by user-friendly tools and resources that could simplify the process of managing salt intake via practical, easy-to-follow steps (particularly for older adults).

Potential intervention recipients highlighted how financial constraints often force individuals to choose cheaper, high-sodium processed foods over healthier options (*Opportunity: Innovation Recipients*). Additionally, limited availability of healthy foods (e.g., food deserts) further confounds efforts to maintain a low-sodium diet. All participants agreed that technology, including health apps like SaltSwitch, could facilitate behavior change. However, challenges such as limited access to smartphones, poor internet connectivity, and usability issues (especially for some older individuals who may not be tech-savvy) were noted.

Potential intervention recipients and RMAs cited the perceived inconvenience of dietary changes, and the effort required to understand and apply nutrition information as major challenges to behavior change (*Motivation: Innovation Recipients*). De-motivating factors for reducing sodium intake included lack of immediate benefits from reducing salt intake, cultural norms, emotional comfort derived from food, and deeply ingrained preferences and routines. To counter these barriers, participants described personal successes in managing their sodium intake and enhancing their diet’s nutritional value by incorporating smoothies and meal replacement shakes, adopting sustainable habits, and making incremental changes. Further, participants emphasized the importance of patience and self-compassion in achieving long-term health goals and healthy eating behaviors.

Healthcare professionals noted that their motivation to address dietary sodium among their patients was affected by the variability in individual biological responses to salt and the perceived lack of control over its effects on BP; thus, they did not tend to emphasize salt reduction unless a patient has persistently high BP despite use of antihypertensive medication (*Motivation: Innovation Deliverers*). They also shared how expressing empathy and sharing personal experiences can inspire healthier habits among their patients.

#### Implementation Process

Implementation Process includes the activities and strategies used to implement COMBI-SR and SaltSwitch in Chicago’s South Side.

To prepare the community for implementation, all participants recommended various forms of education as the first step in promoting healthier lifestyles within their communities (*Planning*). For example, participants suggested integrating health ministries and walking groups into other church activities, researching food sources for a clearer understanding of what community members are eating, and recognizing potential risks associated with certain foods (e.g., contamination, harmful chemicals).

Participants emphasized allowing community members to set achievable health goals (e.g., exercising 3 days/week for 15 minutes) to boost self-efficacy and create a sense of accomplishment (*Tailoring Strategies*). They stressed focusing on long-term well-being, with aspirations like seeing grandchildren as a motivation for thriving rather than just surviving. They also valued visual learning, as demonstrations showing the hidden amounts of salt in foods will make the impact of education on dietary choices more effective. They expressed dissatisfaction with unclear information about sodium and advocated for straightforward messaging to ensure that health information is easily understood and applicable.

Participants emphasized that effective leadership in dietary education and change should start with professionals who can address the psychological aspects of eating habits, such as counselors or behavior specialists, as well as pastors, heads of households, and formal health promotion roles (e.g., nutritionists, trainers, researchers; *Engaging Deliverers*). Participants also acknowledged the value of grassroot efforts and engaging local leaders to initiate a sodium reduction program. Regarding engagement of community members, participants emphasized the value of affordable educational seminars and cooking classes to teach people how to prepare flavorful, low-salt meals, and the importance of one-on-one education to address individual barriers and encourage behavior change (*Engaging Recipients*). Participants suggested greater community engagement (beyond FBOs) through marketing, leveraging opinion leaders, collaborating with local schools and businesses, and creating supportive environments where members hold each other accountable. Additionally, they highlighted the importance of instilling healthy eating habits early via school-based programs.

Participants identified approaches to ensure that sodium reduction becomes internalized, practiced, and shared, to ultimately transforming into a habitual and sustainable lifestyle change, such as: increasing physical activity to naturally reduce salt consumption through an overall healthier lifestyle; receiving personalized health advice from physicians through transparent and direct discussions about health; and modifying diets to recalibrate taste preferences, rendering previously acceptable levels of salt intake excessive (*Doing*).

Participants expressed a desire for simple, accessible changes that make dietary sodium adjustments easier and more effective within their neighborhoods (*Adapting*). For example, given that community members may feel uncomfortable in healthcare settings, community-based programs in familiar environments (e.g., churches) may be a more welcoming environment for implementing a sodium reduction program. Messaging recommendations included using small, actionable messaging (e.g., “shaking the saltshaker less frequently”) rather than abstract measurements (e.g., grams/day).

### Strategies for Implementing COMBI-SR

Implementation strategies refer to the methods or techniques to enhance the implementation and sustainability of an innovation. To successfully implement COMBI-SR in Chicago’s South Side, participants responses were coded into 20 strategies from the ERIC compilation (**Supplemental File 4**).

Healthcare professionals mentioned that an important strategy for implementing COMBI-SR includes continuous refinement of implementation efforts through *evaluative and iterative strategies*, particularly by conducting local needs assessments to collect and analyze data on sodium impact and sharing these findings with key executive leadership in government to gain their support. Participants noted the importance of *adapting and tailoring strategies* to local contexts, recognizing that each community has unique barriers and facilitators that must be addressed to ensure effective sodium reduction. Key strategies include securing buy-in from executive leadership in government, providing personalized, team-based care with nurses and nutritionists to address patients’ unique circumstances, and maintaining continuous patient engagement through individualized education. *Developing strong stakeholder interrelationships* emerged as a key approach, with a particular focus on engaging community leaders, leveraging trusted spaces like churches, and partnering with local organizations such as “Feeding America” to build a coalition to reduce sodium consumption in the community. Participants highlighted that observing successful dietary interventions (e.g., community kitchens) at other sites in the community could provide valuable insights for COMBI-SR implementation. They also emphasized the need for ongoing training and education for key constituents, particularly healthcare professionals, to ensure they are well-equipped to support sodium reduction efforts (*Train and educate stakeholders*). Practical approaches such as offering food samples, conducting dietary recalls, and providing tailored shopping guidance were recommended to empower individuals to make healthier choices.

To *support clinicians*, participants suggested revising professional roles and creating new clinical teams, potentially involving medical assistants, social work students, and nursing students, to enhance team support and improve communication with patients. Recommended strategies for e*ngaging consumers* included community-level education, practical resources, and simple steps (e.g., how to read nutrition labels, avoiding extra salt) would help patients feeling overwhelmed by the abundance of information online. They recommended patient education campaigns, motivational interviewing, and food demonstration classes to enhance patient engagement. Healthcare professionals mentioned the need for funding and policy support, such as allowing WIC benefits to be used at farmers markets, to make healthier food options more accessible and appealing (*utilize financial strategies*).

## Discussion

This study explored the contextual factors in Chicago’s South Side and strategies for implementing COMBI-SR via interviews and focus groups with potential intervention recipients, faith-based leaders, and healthcare professionals. Barriers to implementing COMBI-SR included the complex messaging, mistrust of food labels, difficulties with sodium content interpretation, lack of awareness of the sodium content in common foods and the associated health risks, socioeconomic disparities, high costs of healthier foods, limited access to low-sodium options, deeply rooted cultural dietary practices and potential unwillingness to changing traditional recipes. Potential facilitators for successful implementation of COMBI-SR included leveraging community partnerships, engaging faith-based organizations, and harnessing the power of technology through apps like SaltSwitch. The positive reception of COMBI-SR’s public health messaging and the interest in user-friendly nutrition apps suggest that these tools could play a crucial role in promoting sodium reduction in the community.

Our study aligns with previous research on COMBI-SR^11,13,14^ and highlights the importance of adapting the intervention to the local contexts, addressing specific barriers, and leveraging community strengths to reduce sodium intake. While Land et al.^14^ established and engaged a community advisory committee through a consultative process to identify key messages, communication channels, local champions, and groups for COMBI-SR implementation,^14^ they did not publish the determinants and strategies generated from that engagement. Likewise, Do et al. ^13^ recognized the need for formative work to explore how COMBI-SR could be adapted in other communities,^13^ but did not detail the contextual factors and strategies for adaptation. This study provides a comprehensive overview of the key determinants and strategies for adapting COMBI-SR for an urban, predominantly AA neighborhood.

The next step of this work is to test the effectiveness and implementation of the adapted COMBI-SR on sodium intake and BP of adults with hypertension in the community. The insights from this study are important in shaping the implementation of the adapted COMBI-SR program. By leveraging the SaltSwitch app at the individual level and mass media communication campaigns, community outreach, and partnerships with local healthcare and government organizations at the community level, this multi-level intervention aims to significantly reduce sodium intake among adults with hypertension. Applying the findings, such as building a coalition and policy and funding support, will likely enhance user engagement and adoption, particularly by addressing financial barriers that are common in this community.

This work further emphasizes the multifaceted role of FBOs and the integration of cultural and religious considerations into health promotion strategies. Integrating interventions into familiar, trusted community and cultural contexts, such as FBOs, can improve engagement, while addressing gaps in sodium awareness and nutritional education through community-based programs can empower healthier choices. Moreover, addressing gaps in sodium awareness and nutritional education through community-based programs can empower residents to make informed, healthier choices. Encouraging gradual, manageable sodium changes, alongside offering continuous support, is likely to result in long-term behavioral changes. This approach aligns well with established strategies for effective health promotion, enhancing potential for sustained sodium reduction and overall health improvement in the community.^20^ By addressing these crucial factors, both COMBI-SR and the SaltSwitch app can be more effectively adapted to meet the unique needs of South Side residents.

This study has limitations worth noting. First, different questions were posed to the different participant groups. While this allowed us to leverage the diversity of insights provided by different participant groups, it may have influenced the themes that emerged. Additionally, we did not ask participants to directly link the noted contextual factors with proposed strategies, which could have provided a more comprehensive understanding of the COMBI-SR implementation process.

## Conclusion

Our study identified the challenges and opportunities to implement the COMBI-SR program in Chicago’s South Side. This study engaged potential intervention recipients, community leaders, and healthcare professionals to identify adaptations to program components, potential barriers to program implementation, in a new context, and facilitators to be leveraged. Participants also identified implementation strategies that may address the noted barriers, which were then mapped onto the ERIC compilation. Next steps include conducting a large-scale clinical trial to evaluate the effectiveness and implementation of adapted COMBI-SR in Chicago’s South Side. Our findings highlight the importance of culturally tailored, community-based approaches in health promotion and underscore the need for supportive policies that address the broader social determinants of health.

## Data Availability

All data produced in the present work are contained in the manuscript

## Acknowledgement

The authors wish to thank the participants who took part in this study.

## Author Contributions

Research concept and design: Sanuade, Carroll, Smith; Data analysis and interpretation: Sanuade and Carroll; Manuscript draft: Sanuade, Carroll, Watson, Ye, Hill, Chipman, Wilson, King, Kho, Del Fiol, Davis, Smith; Acquisition: Sanuade, Smith.

## Data Availability

All data produced in the present work are contained in the manuscript.

