## Supplemental File 1 for "Contextual factors and implementation strategies for a multi-level community-based sodium reduction intervention in Chicago’s South Side: A qualitative study"

**Supplemental File 1: Interview Guides**

A. **Intervention Recipients and RMAs**

**Step 1: Registration**

When **registering** for conversation participants share some basic demographics:

Age, race, gender and What is the primary device you use at home to get on the internet? (Prompt - Desktop computer / Laptop computer / Tablet / Phone)

Have you ever had trouble accessing the internet?

i. A member of the faith-based community
ii. A member of the faith-based community concerned about personal salt intake or aware of high intake in the family
iii. A health professional and church member
iv. A volunteer in faith-based community roles such as health ministry
v. A Caregiver of someone in the faith-based community
vi. A Church leader (bishop, elder, pastor, deacon)

**Step 2: Conversation: You and Salt**

TRCDO/P4P is part of a research study group which is interested in hearing from you about the things that are important to your health and wellbeing. Today we are going to talk about Salt and public health messaging about salt what our research partners refer to as Communication for Behavioral Impact for Salt Reduction or (COMBI-SR) for short. We’ll come back to COMBI later!

**Icebreaker (10 mins)**

**Q1: What immediately comes to mind when you think about salt? And What about when you think of sodium?**

*(Prompt - Can you say why you think of that?*

*What or who do you associate that with?)*

**Q2: Does Salt have a specific meaning in the faith-based community?**

(Salt is sometimes described as “the cure for anything (sweat, tears or the sea), hard work honesty. It can describe a very good and honest person or group of people.)

**Q3: Overall, how do you feel about salt?**

*Thanks for sharing. Now we’ll talk about what’s important to you and your community.*

*Participants are invited to respond to questions about their lived experience with salt. Topics are grouped under the following topic categories:*

**Topic 1: Your knowledge about daily salt consumption (15 mins)**

**[if time] Q1: “People don’t know the amount of salt they consume on a daily basis’’ –**

*(Prompt - To what extent do you agree with this statement?*

*If yes – you agree – why do you think that is?*

*If no – you disagree – why do you think that is?)*

**Q2: What does high salt intake look like?**

**Current dietary guidelines for sodium intake in the US recommends adults limit sodium intake to less than 2,300 mg per day—that's equal to about 1 teaspoon of table salt!”**

*(Prompt – where would you put your consumption of salt on a daily basis?*

*does it matter how much salt we consume on a daily basis?)*

**Topic 2: You and salt management (15 mins)**

**Q1: Salt intake cannot be controlled**

*(Prompt - To what extent do you agree with this statement?*

*If yes – you agree – why do you think that is?*

*If no – you disagree – why do you think that is?)*

**Q2: How much would you say you know about managing your salt intake?**

(*Prompt If a lot – where do you think you got that knowledge and understanding from?)*

*(Prompt if not much – what do you believe are the gaps in your knowledge or understanding?)*

**Q3: What helps you to manage your salt consumption?**

*(Prompt - food labeling of sodium; recipe info; choosing foods relatively low in sodium, consciously limiting the amount of salt added in food preparation and at the table	identifying the major foods that are low in sodium)*

**Q4: What activities help you to consume less salt?**

*(Prompt – cooking with only small amounts of salt, avoid adding salt at the table, avoid salty prepared foods, purchasing foods with reduced salt and sodium content; Manufacturers of processed foods reducing sodium by 20% from present levels and advertising salt reduction)*

**Topic 3: Your public health messaging and social media (20 mins)**

**Q1: When you see messages like this – how do you respond?**

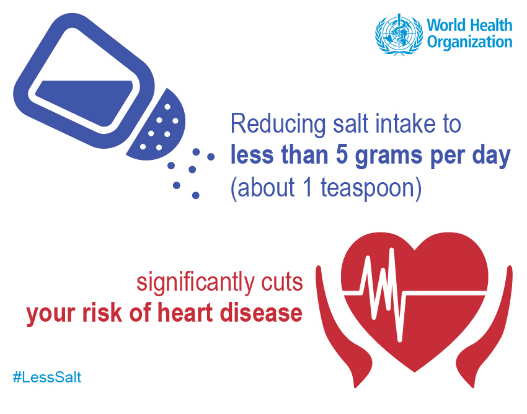

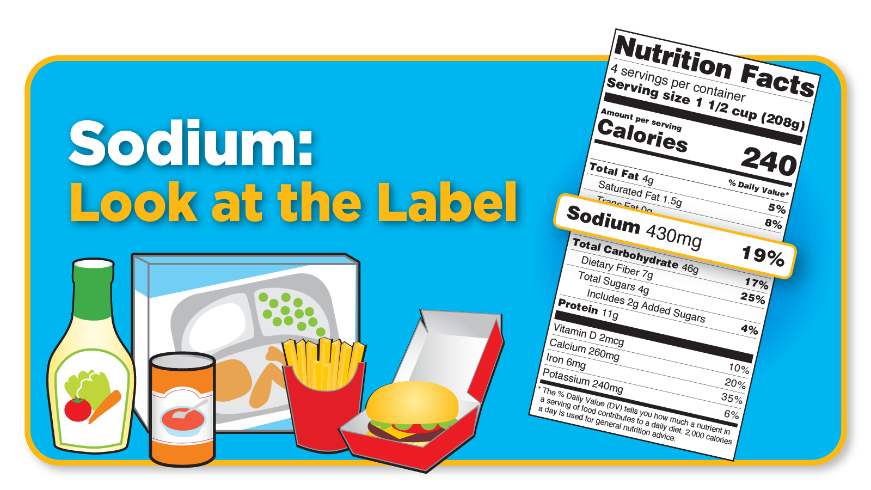

**Q2: Can you tell me about the media or technology platforms you have used for health and wellbeing?**

*(Prompt – Online support groups Facebook /Linkedin / Health Apps/ Fitbit type technology/ Telehealth / Television/Radio/Newspaper/I do not use media or technology for health and wellbeing Other☺*

**Q3: If there was a health app that could provide users with easy access to nutrition information, including salt levels, along with a list of lower-salt alternatives to which they may ‘switch’ how likely would you be to use this?** Why or why not?

**Topic 4: You and your community (15 mins)**

**Q1: What types of salt or sodium do you and your family use for cooking or meal preparation?**

*(Prompts e.g. rock salt; kosher salt; sea salt; salt flakes) Prompts: do you drink or use salty liquid versions like chicken or vegetable stock?)*

**Q2: Do you think people in the community consume too much salt?**

*Prompt: Any particular reasons you would like to highlight?*

**Q3:** **What do you think are the consequences to the community of consuming too much salt?**

**Q4: In your opinion, are there certain** **places or services in your community you can you use to manage salt intake? (e.g. grocery stores; pharmacy)**

**Q5: And what activities can help members of your community consume less salt?**

*(Prompt – examples of cooking with only small amounts of salt, avoid adding salt at the table, avoid salty prepared foods, purchasing foods with reduced salt and sodium content; Manufacturers of processed foods reducing sodium by 20% from present levels and advertising salt reduction)*

**Q6: Who or which organization(s) should be involved in connecting community members to community resources for salt intake management?**

**Addendum (RMAs only)**

At the start of this interview I mentioned the name of the research study sub group – COMBI SR and explained that this stands for Communication for Behavioral Impact Salt Reduction (COMBI-SR) .

**Icebreaker:** What do you think COMBI-SR researchers might be focused on?

Ok thank you! So one thing the COMBI researchers are doing is combing through the published research literature to select activities to help everyone reduce salt intake . One activity which people are using in countries around the world (e.g. including UK, New Zealand and Australia) – is something called a salt switch. Take a brief look at this picture of the Salt Switch App version.

(Prompt: *What do you see?)*

**Q1A: So, in your role as an ambassador, would you feel comfortable encouraging members of your faith-based community to use an app like this to manage salt intake?**

(*Prompt If yes – what additional knowledge and understanding might you need?) Prompt if not – what do you believe are the barriers to use? E.g., do you know enough about salt levels, to make the ‘switch.’ Yourself to a lower salt alternative?*

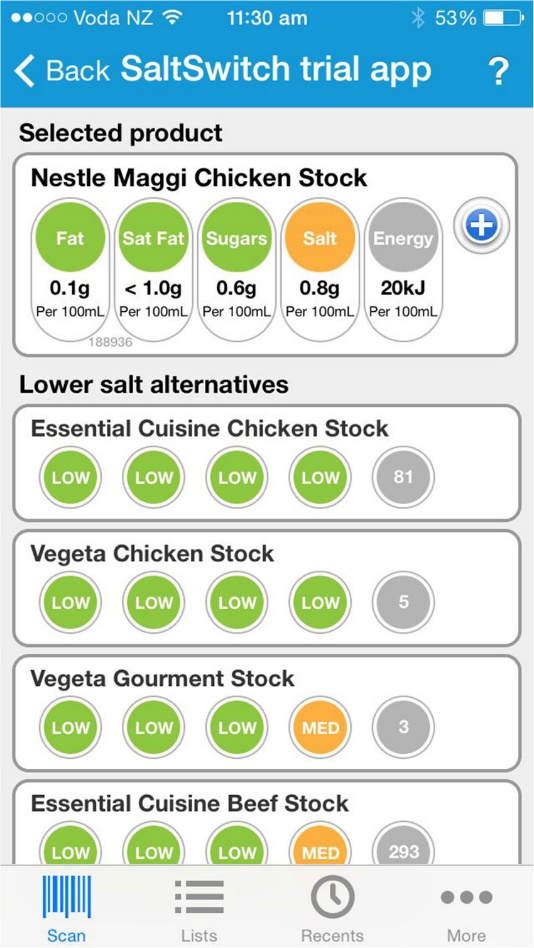
**Q2A: Have you or your faith-based organization advocated for dietary change like salt reduction before?**

*(Prompt If so can you explain how this was done; and what the outcome was? Any lessons learned?)*

**Q3A: Are there people in your organization who can champion salt reduction activities?**

*(Prompt If yes – who might they be; the skills they have: If no, who might deliver?)*

**Q4A: What additional faith-based community activities on how to reduce salt intake might help community consumers to switch away from the saltiest products when they go shopping?**

*(Prompt: education provided by an external provider, communications, other targeted events demos)*

**Q5A: How do you think the individuals served by your faith based organization will respond to the organization leading the invitation to reduce salt?**

*(Prompt: What do you think might be some of the barriers or challenges facing faith based community members trying to switch to healthier yet similar salt alternatives? (E.g. Cost of alternatives?; Having to abandon favorite brands; there are no healthy alternatives; no phone to host app )*

**Q6A: What might your organization need to be able to implement and deliver activities like these?**

(*Prompt: the sky is blue at this point – would you need for example* ***meeting support*** *- arranging group meetings, partnership sessions, school activities, cultural events, leaflets, posters, pamphlets, videos, and home visits on importance of reducing salt intake and other practical ways to achieve a reduction. Or* ***support for advertising*** *- e.g. about the importance of salt reduction and practical ways to achieve this reduction.* ***Or resource support*** *: e.g. information booths in churches on how to reduce salt intake ; education on how to use the Salt Switch App when shopping for household food and beverages to increase selection of low-salt foods. Point-of-service/sale promotion information on local stores /pharmacies where people can get salt substitutes and providing salt substitutes to enrolled participants at no cost)*

**Q7A: Any other comments you would like to share at this time?**

Thank You so much for your time today it is very much appreciated. Your thoughts and insights are an essential component of making the Salt Switch!

B. **Health Professionals**

**Barriers and Facilitators to Reducing Salt Intake**

1. To what extent will an intervention to reducing salt intake be welcomed in the patient population that you serve?
2. What are the barriers, from your perspective (as a clinical team, provider, etc.) to reducing salt intake in your patient population/community?

***Prompts:***

- *How much of a barrier is cost of low-sodium foods for your patient population?*

3. How do you think these barriers can be addressed?
4. What roles do you think healthcare providers can play in addressing excess salt consumption in your patient population/community?

From your perspective, what are the components of an effective and sustainable approach to reducing salt intake at the individual, family, community level?
