## Supplemental File 2 for "Contextual factors and implementation strategies for a multi-level community-based sodium reduction intervention in Chicago’s South Side: A qualitative study"

**Supplemental Table 1. Codes Applied to Consolidated Framework for Implementation Research (CFIR) and Expert Recommendation for Implementing Change (ERIC)**

| **CFIR DOMAIN - CONSTRUCT (N=532)** | **N quotes** | **% (within domain)** |
| --- | --- | --- |
| **INNOVATION** |  |  |
| Relative advantage | 22 | 37.9% |
| Cost | 7 | 12.1% |
| Design | 12 | 20.7% |
| Complexity | 8 | 13.8% |
| Evidence-base | 9 | 15.5% |
| **OUTER SETTING** |  |  |
| Local attitudes | 35 | 33.0% |
| Local conditions | 40 | 37.7% |
| External pressure | 1 | 0.9% |
| Financing | 1 | 0.9% |
| Policies & laws | 4 | 3.8% |
| Partnerships and connections | 25 | 23.6% |
| **INNER SETTING** |  |  |
| Access to knowledge and information | 65 | 47.8% |
| Available resources | 19 | 14.0% |
| Mission alignment | 2 | 1.5% |
| Available resources: Funding | 4 | 2.9% |
| Available resources: Materials & equipment | 1 | 0.7% |
| Communications | 7 | 5.1% |
| Compatibility | 7 | 5.1% |
| Culture | 13 | 9.6% |
| Structural characteristics: Physical infrastructure | 5 | 3.7% |
| Structural characteristics: Work infrastructure | 1 | 0.7% |
| Relative priority | 12 | 8.8% |
| **INDIVIDUALS** |  |  |
| Motivation | 56 | 29.3% |
| Opportunity | 21 | 11.0% |
| Capability | 78 | 40.8% |
| Need | 36 | 18.8% |
| **IMPLEMENTATION PROCESS** |  |  |
| Adapting | 3 | 7.3% |
| Doing | 5 | 12.2% |
| Reflecting and adapting: Implementation | 1 | 2.4% |
| Engaging: Innovation deliverers | 6 | 14.6% |
| Engaging: Innovation recipients | 18 | 43.9% |
| Planning | 2 | 4.9% |
| Tailoring strategies | 6 | 14.6% |

| **ERIC Codes Applied (N=75)** | **N quotes** | **% (within domain)** |
| --- | --- | --- |
| **1: USE EVALUATIVE AND ITERATIVE STRATEGIES** |  |  |
| 1.7. Conduct local need assessment | 3 | 100.0% |
| **3: ADAPT AND TAILOR TO CONTENT** |  |  |
| 3.1. Tailor strategies | 1 | 25.0% |
| 3.2. Promote adaptability | 3 | 75.0% |
| **4: DEVELOP STAKEHOLDER INTERRELATIONSHIPS** |  |  |
| 4.1. Identify and prepare champions | 3 | 18.8% |
| 4.5. Build a coalition | 9 | 56.3% |
| 4.9. Capture and share local knowledge | 4 | 25.0% |
| **5: TRAIN AND EDUCATE STAKEHOLDERS** |  |  |
| 5.1. Conduct ongoing training | 2 | 22.2% |
| 5.7. Conduct educational meetings | 7 | 77.8% |
| **6: SUPPORT CLINICIANS** |  |  |
| 6.4. Revise professional roles | 1 | 25.0% |
| 6.5. Create new clinical teams | 3 | 75.0% |
| **7: ENGAGE CONSUMERS** |  |  |
| 7.1. Involve patients/consumers and family members | 7 | 20.0% |
| 7.2. Intervene with patients/consumers to enhance uptake and adherence | 14 | 40.0% |
| 7.3. Prepare patients/consumers to be active participants | 13 | 37.1% |
| 7.5. Use mass media | 1 | 2.9% |
| **8: UTILIZE FINANCIAL STRATEGIES** |  |  |
| 8.1. Fund and contract for the clinical innovation | 1 | 25.0% |
| 8.2. Access new funding | 3 | 75.0% |
