## Supplemental File 3 for "Contextual factors and implementation strategies for a multi-level community-based sodium reduction intervention in Chicago’s South Side: A qualitative study"

**Supplemental Table 2. Barriers and facilitators to implementing the COMBI-SR program, a multi-level community-based sodium reduction intervention, in Chicago’s South Side community**

| **CFIR Domain and Construct** | **Definition** | **Theme [Barrier/Facilitator]** | **Illustrative quotes** |
| --- | --- | --- | --- |
| Innovation |  |  |  |
| Overall context of sodium consumption | Perception on what immediately comes to the mind of participants when they think of sodium. | Causes disease (high blood pressure, headache, body swelling, edema). | The first thing that comes to mind for me (when I think about salt) is high blood pressure. (RMA 5, IDI)  When I think of sodium... I just automatically think bad, not healthy for my body need to find ways to minimize it. So I equate salt sodium, all as bad. (PIR 3, IDI)  … depending on what type of salt. Some salt can cure headaches, some salt can cause them (PIR, FG) |
|  |  | Processed/junk/fried foods. | Maybe a lot of fried foods **have** salt like, I think, about chicken like hot sauce. A lot of sauces. Junk food like chips. not so much as candy, but definitely like over processed foods. (RMA 4, IDI). |
|  |  | Cure for headache | … depending on what type of salt. Some salt can cure headaches, some salt can cause them (PIR, FG) |
|  |  | Flavor/seasoning/taste enhancer. | That's a good point. I didn't even realize about the sodium being a nutrient. For me when I think of salt, I think about taste like I could just see somebody like maybe putting it on some sort of meat or something. But, like, you know, really laying it on or trying to enhance the flavor of something. (PIR, FG).  Just jumping off both of you guys this point. And I think of salt, I just think of flavor this kind of the go-to, for, like all things seasoning if you want anything to taste good, you put salt on it. That's just kind of always been the rule of thumb especially in my household. (PIR, FG). |
|  |  | Faith-related (punishment, salt of the earth). | So that's what comes to my mind. I may not know the entire Bible, but there is a story in there about a lady who turns into a pillar of salt. So salt was a destiny you know, a death sentence to her, and I think it was because she was standing by her husband and lied about something. You know what I mean, or something like that. So in the Biblical sense, it's not used for seasoning its use is I don't wanna say punishment, but it could be used in a lethal way. Let's say that a lethal way (RMA 2, IDI). |
|  |  | Energy enhancer | When I think of sodium - since I'm really into fitness - I think of like the healthier side of salt intake, because you need sodium like to work out to have energy (PIR, FBD). |
| Innovation Relative Advantage | Perception that SaltSwitch app is better than other available innovations or apps. | Concerns about suitability for weight loss and potential product alternatives. | How is this? Gonna help me? I'm trying to think of some other questions. You know, something a lot of people say, is is this gonna help me lose weight, like, you know, How is it gonna help me if I use this app? And I've just heard that you say, you know when you go to the store. It give me a better alternative of what product I should buy instead of that one you know. So with the having less salt intake Is it going to refer me to something that would taste good?. Or you know how many people are actually using this product that you suggest, and [why] I should change to you know, how many stars is it, that kind of thing? (RMA 5, IDI) |
|  |  | Competing dietary restriction programs and fitness/exercise apps/social media groups (e.g. peloton, Nike running app, iphone fitness app, PAL, Yuka, Fitbit, Youtube, Facebook, Twitter, DVDs) | I have a peloton at home. and so I use the app for that. I also use the what is it called? The Nike running app I've used what is it called? Yeah, the fitness app on my iphone. And there's this app called run keeper that I used to use. Youtube videos here and there. But that's for exercise. Yeah.(PIR 7, IDI)  So I don't know it's like this, app the Yuka app, and you can like scan, bar codes and everything like at the general store, and I'll have like you can use it with, not just like blue products. You can also use it with like aircraft products and everything - literally go to like the ingredients and kind of tell you if it's excellent low risk. A friend showed me this, and you'd be kind of surprised because they'll tell you like all those head head boggling like words. If I call those chemicals, it'll just like, even if you don't know it. It'll just tell you like, Hey, this is not like this is a poor quality ingredient, so that at least, even if you don't know or have the time to do the research on that ye most people will jump at the fact of like Oh, I this is a poor quality thing, and go to a better alternative and say, just kind of going in blind or learning the hard way after usage so (PIR, FG)  Well, a friend of ours started a Facebook group that we are part of called. It's a wrap. And it's like it's just overall wellness. And it's like motivation. She'll show herself at the park or at the track, or just her workout. She will show, you know, I mean calories and kind of like a motivation. Also, just trying to hang with my young adult kids, you know, they always doing something and so I get out there with them. Try to walk, walk with them, you know. Incorporate some stuff in my lifestyle that will enable me to have fun. Monitor my weight, and just try to have a healthier lifestyle, you know. (RMA 1, IDI) |
|  |  | Potential use of SaltSwitch if it is free and affordable | It's something that I feel like I will use. But honestly, I feel like there's already apps on the market like that. I'm just too cheap to purchase them. So if it were free and you know readily available, I would be more apt to use it but the older I get, the more I'm realizing that we have to invest in our health. no matter how that looks, because our health is the principal thing. It's our, the primary thing, like we. We want to live long and healthy and prosperous lives. you know, not only economically, but but also, as far as health is concerned. so yes, I would absolutely be interested in it, even if I had to pay for it. And again, I think there are apps I need to explore already that will allow you to scan and tell you how much of something is in, isn't it? But yes, sir, I think that I believe that I would and support it. Yes.(PIR 6, IDI) |
| Innovation Complexity | Perception that the COMBI-SR/SaltSwitch app is complex, possibly due to its extensive scope, numerous features, or the multiple steps required for use. | Confusion over nutritional labels and serving sizes | So, being health conscious, I often wonder, ‘How much salt is in this bag of chips? How much salt is in this item?’ The way nutritional information is written is confusing. You might think the listed amount is for the whole item, but it’s often for a single portion. A box might have 7 or 8 portions, so you have to multiply to figure out the total salt content (PIR 5, IDI) |
|  |  | Lack of awareness and understanding of nutritional labels | Some people just look at the label. I don't know if they were reading it, you know. But I can't really look at this and know - 4,500 grams of sodium in it. Do you know how much that is, you know. Some people just not even aware of it, of how much salt in or sugar is in things that they eat. (RMA 3, IDI) |
|  |  | Challenges in implementing nutrition education at the store level | They wouldn't let you do that because you couldn't put a form to say, read your labels. You're only supposed to have less than 2,300 grams of salt. That's because they feel that stops people from buying certain products. So I don't think there's anything you could do unless they let you, which I don't see happening. Like how they have wine tasting—you know, they would have a person in the store to show you. But I can't see what you could do at the store level—to me, it's gotta be a personal thing (RMA 3, IDI) |
|  |  | Complexity of the app | the creators or builders of the app to make sure that they include that type of information in the app . Make it easily accessible for the, you know, because the people can't - like they don't fool around like I did with the zoom thing today. They'll be like, Okay, I just can't do it. It's gotta be something they can access easily. And it be just plain, simple language. You know things that everybody can understand, and in a way that they can understand. Just like you just said, Okay, if I had a bowl of spaghetti and I put 2 teaspoons of salt in it, that's too much salt. So I mean, so that I could understand. So some things they maybe have to break the measurements down. you know, right? It was like when my mom was a salt user, and we had to like hide the salt from her. So I found if you can try to get people to understand, you know that this might taste good to you right now, but in the long run it's damaging your body. But I guess they couldn't say that that ain't gonna make 'em use it. But mostly I think plain and simple instructions, and plain and simple log in to get into it. You know some people get real excited about new apps. You know.information like that, I think, would make it more usable. (RMA3) |
| Innovation Design | Perception that COMBI-SR/Salt Switch app is well designed and packaged, including how it is assembled, bundled, and presented. | Appreciation for detailed nutritional information | And I really love the screen you showed when you could scan something on your phone. It shows all the content of the sodium. And this, that and the other. So, yeah, that's that's that's huge. That's huge. So I'm looking forward to the next step (RMA 1, IDI) |
|  |  | Enhancing saltswitch to provide alternative product recommendations and purchase locations | In the moment it might be good. I think Yuka does I think it might I don't know. Sometimes it goes off store. So you're in the store. These are like the better products here. It would need gto be developed more like telling you where to go; like what's the next best thing. But for like a startup, I think it's a good for the benefits later. (PIR 4, IDI) |
| Innovation Cost | Perception regarding adoption of COMBI-SR/SaltSwitch app if affordable. | Adoption potential depends on SaltSwitch being free | Oh. honestly, if it were free – yes (I would use it-SaltSwitch). I don't know if I would pay for it. (PIR 7, IDI)  I think people of all demographics like the word free. So if there isn't a cap where you can provide services for people to get them to get them going not to be a crutch, though - cause we don't wanna baby nobody for the rest of the walk in life. But at the same token, if the resources are available, we can get you can. We can get very creative with it. And you know, have ha! (RMA 1, IDI) |
| Innovation evidence-based | Perception on the challenges and need for further research to accurately understand the biological effects of salt consumption, beyond current hypotheses and anecdotal assumptions. | Complexity and uncertainty in understanding salt's biological impact | so like I said, for me, in my opinion, I still think that, as it relates to biology as it relates to food, we still need a hundred more, a hundred to 400 more years to really speak accurately about like hat salt is really doing in someone's body, instead of just going with the hypothesis like, Oh, this person eats too much fried chicken with salt , and that's the reason why they developed this (PIR 4, IDI) |
|  |  | Concerns about how (daily) recommended sodium intake guidelines are established and their relevance to varying levels of physical activity and fitness routines. | I think this is more of a question, maybe for the both of you, because you're researching it. Is that number just for the person who lives like on average, like the average person, as I know when me and FG4, the point we have made earlier, like we like ran track, and in some part of our lives, and like me as somebody who's passionate about fitness like have a pretty intense like workout regiment. So generally I have to consume more salts, cause that's like what's recommended because you're sweating so much. You're like losing salt and having too much salt is bad, but also having too little like sodium in your body, especially when you have an intense fitness, regiment is bad for you, and then can cause other issues. So I was wondering [how that number was reached] like what factors were taken into account about like lifestyle, or is that on average I was just more curious about where the number came from. (PIR, FG) |
| Outer Setting |  |  |  |
| Local attitudes | Focuses on how salt symbolizes preservation, transformation, and spiritual responsibility within faith-based communities. | Symbolic role of salt in faith-based communities | When it comes to faith, faith-based communities, I do. I believe that salt, I mean, if they're saying, I mean what Scripture says, I believe that 'ye are the salt of the earth.' And if that is the case, and if we are the salt of the earth, then there should be some kind of, I think, as far as faith-based communities are concerned, some kind of preservation or, yeah, some kind of preservation that should be accompanied with, you know, being a Christian and carrying the banner of Christ. If that makes sense (PIR 6, IDI) |
|  | This theme emphasizes the role of salt as a symbol of togetherness and how seasoning with salt makes food enjoyable and comforting, fostering gatherings and communal meals. | Salt as a symbol of communal bonding and nourishment | Salt, to me, symbolizes coming together and family. When it's sprinkled on food, it enhances the taste and brings comfort, often drawing more people to gather around during events, gatherings, or meals. Biblically, I see it as a metaphorical meeting place, signifying a place of connection and community (RMA 1) |
|  | This theme highlights how use of salt is pervasive in daily food choices, festive occasions, often unaware by people, demonstrating how seemingly harmless eating habits can result in substantial cumulative sodium intake, potentially affecting long-term health | Lack of awareness of sodium intake from different sources | I believe people often overlook how much salt is in these foods, beyond just using salt to taste. This can be problematic because your palate might be accustomed to high salt levels without realizing it. The lack of clarity about salt content on packaging makes things worse. Some foods intentionally have too much salt, and others have hidden salt levels that consumers aren't aware of. For example, bacon and turkey bacon both contain significant sodium, and even turkey burgers have some sodium, despite being a leaner choice. (PIR 4)  You know, I'm really surprised, and let me explain why. I feel like even my toothpaste has salt in it. It makes me wonder what's really going on. Are we being set up for challenges in our communities and lives? It feels like we might have been sold a lot. It's like there's something deeper at play here, because we're told we should only have one teaspoon of salt... who even realizes this? (PIR, FG)  I don't believe that most people are measuring what they ingest like the amount of sodium or salt that they're ingesting daily, whether it's like cooking their food, or even like in their beverages. And especially, I think it's borderline impossible to measure it if you eat out. So that's why I would say it. I don't think most are doing that. (PIR 7) |
|  | This theme shows individual’s awareness of high sodium in packaged foods and their deliberate efforts to manage intake of these foods. It reflects a balance between health awareness and personal culinary choices in salt usage. | Awareness of hidden sodium in foods | Based on what I see on these packages, some of the foods are like, 'Okay, I wanna buy something that's quick and convenient.' But when I look at the back of the package, it's a label, you know. You see all these, you know 300—I don't know if it's milligrams—all these grams of high sodium. You know it looks good and may taste good, but just thinking about eating all that down the road, it can, you know, again, affect the blood pressure that I really care about. It's been doing well with. And I really wanna undo the process of having medication. So I really try not to use salt at all. I, you know, like the Morton salt. I buy it, but I actually use it in the wintertime, for the steps. That's how strong salt it is, it could break ice. It could break snow, you know. You put them on your stairs. I mean, it's cheap but that's good for your stairs. (RMA 5) |
|  | This theme highlights the socioeconomic disparities in access to healthier, low-sodium foods. It underscores the challenges faced by communities without access to higher-end grocery stores, where cheaper canned and processed foods often contain higher sodium levels. | Socioeconomic disparities in access to low-sodium foods | " I do believe. And I do believe, because in the hood, there is only a certain amount of fresh produce and fresh foods and fresh meats and things that you're able to get. So, a lot of people in the hood are buying what is economically feasible for the situation, and because of proximity to not having the option for fresher items or items with less preservatives. Yes. I do believe that the salt intake is more." (PIR 6) |
|  | This theme emphasizes how hectic schedules prompt many individuals to choose processed, preservative-rich foods with high sodium levels. It also discusses the prevalence of fast-food culture, known for its affordability and convenience despite its high sodium content. | Societal dynamics and easy access to fast foods | We eat a lot of fast foods and all that's nothing but salt, unhealthy. That's the major thing, you know. I remember when I was in college, we used to say, you know, and you're young. So you're joking. But at the practice one thing about, you know so so, banana we like. We want a brown paper bag with grease at the bottom. That's quick, cause you're too tired to cook. So that's the world we live in, but a lot of times. It's truly not. It's not healthy. It's not healthy for us at all. Now there are some. I'm not gonna bad every place that you can get food in 2 min from a drive through.(RMA 1)  ..I don't know if civilization is the right word … But the way our society works kind of works against us, and I'll say most people don't know how much sodium they're intaking because they don't prepare their foods on their own. We live in a highly grab and go society, or we're looking to cut corners or be more efficient. So if I have to focus my energy on work, or if I have to focus my energy on my family, I'm looking for easy ways to like make food. So even if I do go to the grocery store, I'm probably gonna grab something that maybe can last for a long time. It has a lot of preservatives in it which will probably have more sodium in it, so that I can, you know, have it last for longer than a week or 2. A lot of People complain now, even when they get food from the grocery store that it's not lasting as long. So then I feel like that ends up leading to us, you know, just getting more stuff that's on the inside of the grocery store instead of on the perimeter, like that sort of thing, and then I can't even speak to fast food. Of course we have a huge fast-food culture. It has to be full of sodium. So I think I'm shocked. And I'm also just thinking to the point that it's almost working against us, us, just trying to make it through our lives and eat on the way. (PIR, FG) |
|  | This theme highlights the cultural significance of food practices across generations, alongside their evolving health implications in contemporary society. | Cultural significance of foods and seasonings across generation | " I think because we season our food more than any other...  I believe we season our food, and I think over time, generational, it became just a habit, not even tasting the food to see if it tastes good, we automatically always add , and I think that over time we're not as healthy. We don't do the same thing. We're not outside as much. So. All that sodium over, the years passed down from your grandmother and uncle, and your and our ancestors. I think it caught up with us now, cause we're not out. We're not working out. We're not. We're not walking out running. We're not drinking enough water, so it didn't affect us as much, I don't think, save my grandparents, but it affects us now because we don't get out as much to counteract all the other stuff we put in our bodies that could be hurtful to us." (RMA 1) |
| Local Conditions |  | Barriers to adopting healthy alternatives due to cost | if I would say, probably it may be difficult - but providing other options or cheaper options for healthier meals and less fast-food restaurants that are in our neighborhoods. I think. Yeah, I think that contributes a lot of it. And so maybe the the fast-food restaurants can provide - well, they do provide alternatives - but I think cost issues is just the main thing of like knowing. (PIR 8, IDI)  I agree. It's really about a complete shift in mindset. A big part of it is the cost. The cheap foods out there taste so good, you know? If you're on a fixed income and using a food assistance card, you can get a lot more unhealthy food for your family than if you were buying healthier options. Healthy food tends to be more expensive. So, I think those are the main barriers, along with education. (HP, FG 2) |
|  | This theme explores the challenges individuals face when considering healthier food options due to lack of knowledge about them. | Barriers to adopting healthy alternatives due to lack of knowledge | There is no food places, not even McDonalds. So this isn't just a full desert for anyone who don't have a transportation. It's almost impossible to buy food from outside unless they cook or food of their own, you know. So, for people with lot of choices, you know, we can say, you know, who can cook at home, who can cut down on the salt. That would be great idea. But people who are eating out all the time, or eating unhealthy food. I don't know if that intervention will be, it's great to suggest. But how practically it will be will be something I have serious doubts about. (HP, FG 2) |
|  | This theme explores the challenges individuals face in choosing healthy foods due to comfort and familiarity with certain unhealthy foods such as McDonald’s French fries | Barriers to adopting healthy alternatives due to familiarity with unhealthy foods | I think the barriers are the cost of healthy alternatives, lack of knowledge about them, and comfort with familiar, less healthy options. For example, what could replace McDonald's French fries in terms of taste and satisfaction? These challenges make it hard to choose healthier foods. (HP, FG 2) |
|  | This theme highlights that while food banks provide shelf-stable products, these items are often unhealthy. It also points out the disparity between larger food depositories and smaller community-based food banks, where donation quality varies widely. | Challenges with nutritional quality of Food Bank donations | I think one of the issues with foodbank is they can provide shelf stable products, right? But a lot of that is not necessarily healthy. Like I think they do the best they can. Like it's one thing to have like a like, you know, a greater Chicago Food depository versus like a church, you know, like sometimes our patients will just go to the church food bank and it's whatever people donate, right? (HP, FG 2) |
|  |  | Pervasive use of sodium in unexpected products | You know, I'm so surprised, and let me tell you why? Because I feel like my toothpaste. Got salt in it. So what's going on? I'll be being set up for failure in our communities, in our lives. Have we been sold a lot? That's what I'm going is is hidden a deeper nerve, because we supposed to have one teaspoon … who noticed .. I don't recall hearing this in biology. Would it be biology? Some type of science class after like, or health or nutrition? I don't remember hearing this now. I wasn't listening because I went to school to make friends. That's why I'm here today so that I can listen more. And if we could get this record. but what I'm saying is. Yes, that is news to me, and I want to know how do you decide? How do you … are we eating too much like me personally like I said. My back is big. I know I eat too much. (HP, FG 2) |
|  | This theme highlights the importance of leveraging various community resources to promote healthier eating habits and lifestyles. It emphasizes the role of community health workers, schools, churches, shelters, food pantries, community kitchens, and grocery stores as centers for health education and food demonstrations, with a focus on culture and community-centered educational activities. | Role of community engagement and education in promoting healthier lifestyles | I would say doctor's offices for sure, although it can be challenging since they serve patients from all over. Maybe it could affect their pricing too, although I'm not entirely sure. It might not be beneficial for these corporations, which might explain why they don't do it. But I was thinking about grocery stores—they could definitely help. It seems like they already promote other options. Fast food restaurants could also play a role in that regard. As for jobs in general, I wonder if they could do something similar. I'm not sure what a standard process would look like, but schools—elementary, middle, high schools, and even colleges—could definitely help implement better activities for kids. They could be part of the team that makes a difference. (PIR 8, IDI)  But in terms of how to operationalize cooking classes, there are some health centers that have kitchens that can be tapped into as local spaces. Chicago Family Health Center, where I used to work on the South Side, had a community kitchen. So, to identify spaces that have kitchens that you can then use for the cooking demonstrations. (HP, FG 1) |
| Partnerships & Connections | Integrate promotion and distribution of healthier food options into existing support systems and services to reach underserved population. | Integration into exisitng services like shelter and direct food donations | It could be something that we, well, I don't know we could provide to shelters and just kind of sneak it in and encourage them to try it. Most of the time they're going to reach for, they don't want to think about those things. They want to think about how do I get housing or how can I get my life on track? And you know what I mean? It's hard for them. But if you just provide it and just kind of suggest, hey, try this, you know, um, try this instead. Maybe that would work. I don't know. (HP, FG 2).  And then also we get donations of healthy green leafy vegetables and fruits and vegetables doing certain fruit store, I mean full store, which we donate to our patients, too. So those, I think, and things can really help for them to be more eat, more healthy food. (HP 2, IDI) |
|  | Role of volunteers and organizations in distributing fresh produce and healthy foods | Food pantry distribution | I get [to volunteer with] the food pantry, so we give out fresh produce. Which is something a lot of are not doing .. we give out Kang's as well, but like we're, you know, giving out fresh cabbage, giving out, you know corn potatoes. We're giving out meat, so people are just not eating like process stuff in the bag. Some people in my community are eating like worthwhile, 'm like y'all got to. Like, I think, I think we're trending down.(RMA 4, IDI) |
|  | Partnerships with businesses and organizations to increase access to healthy food and education | Partnerships with businesses and organizations | ...even like you mentioned partnering with jewels. Imagine every time they shop they're able to get you know, some type of free fruit or vegetable with a note attached to adjust with some form of simple education. So that's that's how I see it. (HP, FG 3). |
|  | This theme discusses the role of schools and community centers in spreading nutritional knowledge and encouraging healthier habits. | Partnerships with educational institutions and communtiy centers | The school system would be the quickest way. Because children are impressionable, like you know, they kind of soak up anything that they learn, and if it was such a mandate or a standard in the school system I think that that will kinda trickle over into the home and then the child is able to kind of influence the parent. So I think that the school system would be a good catalyst to bring about changes it pertains to. [So to your question] what organizations are getting that information out there for the people .. I'm a going with the school system and community centers. The only thing is who really has a community center? You know, there's not many community centers that I do see. So that's why, again, I'll say the school. But School Community Center, Y.M.C.A., organizations like that. I think that they could play a really important role in getting that health conscious information to the people. (PIR 6, IDI) |
|  | This theme focuses on leveraging trusted community spaces like FBOs to disseminate health information and conduct educational activities. | Partnerships with faith-based and community organizations | Um, training, like if there could be trainings in some of the food pantries or like Catholic Charities when they do their meals, the meal sites, that's the word I'm looking for. Inspiration cafe, different places where people go to get food to work with the people who are cooking to have there be food demonstrations in real time, going to churches. Um, the South side has the highest net density, like more churches than any other community in the US per population. So going into the churches which are trusted spaces to work with pastors, to work with church kitchens, thinking about where are the places where people come together to cook together. Um, and letting there be educational sessions. But really thinking about where the information begins to come from, like once the decision is made, let's do food as medicine. Let's do salt reduction as a medicine within the food that people are taking. The main distributors of food knowledge would be where communities come together to eat and figuring out where those spaces are. (HP, FG 1). |
|  | This theme focuses on the involvement of healthcare providers in educating patient about nutrition and managing dietary intake. | Partnerships with healthcare settings and medical professionals | I would say, doctor's offices for sure. I mean it can be difficult, because I'm sure they have patients from all over. But maybe I mean, this might affect their pricing too. I think it might not. Actually, it might not be beneficial for these corporations. That's probably why they don't do it. (PIR 8, IDI)  The free clinics that we have around here, the doctor's office, the urgent care, the hospital, I think you know is just kind of one of those messages that needs to get out, you know. The same way they they went through all of this about e-cigarettes, and you know what I'm saying. (RMA 2, IDI) |
|  | This theme highlights using community kitchens and gardening projects that engage individuals in healthy eating practices. | Making use of community kitchens and gardens | But in terms of how to operationalize cooking classes, there are some health centers that have kitchens that can be tapped into as local spaces. Chicago Family Health Center, where I used to work on the South Side, had a community kitchen. So to identify spaces that have kitchens that you can then use for the cooking demonstrations. (HP, FG 1).  One more thing. The community gardens too. So community gardens, school gardens, all that growing food. So if you want to even take it a step back (there is a backup plan to continue the initiative). (HP, FG 1) |
| Policies & Laws | This theme highlights need for increased government and policy support, as well as policy and funding sustainability. | Policy and government involvement | …I think that any governmental entity, if you're being paid by a city or a state like this should be number one priority right now outside of like the fact that you know people are getting shot, or you know the poor doesn't have food. But it's like, if the poor are also eating Mcdonald's, then we still have the same issue. So I think that this has to start not just in a household level, but I think, like the State, and the Government has to be way more committed to the the health of our people, and obviously because they are there are already like contingencies and contracts in place is gonna be extremely hard for, like us to figure out like, whose responsibility is it. Because the government is going to say, Well, that's the company's responsibility and then the company is gonna say, well, if the government ain't gonna put no sanctions on us we gonna do, we wanna do. And then the company is gonna say, well, the government allowed it. So it's gonna always be these checks and balances that these dances that's gonna happen. But I do think that the companies that are providing us the goods, and the government should be actually educating us on this. But because it's a capitalist society, it's hard to do that like we're not a Socialist society. So it's kinda like asking a capitalist, a capitalist society to care about people in order for them to lose money .. which is gonna ultimately cause them to lose money. That's really what it is. So II think the First Level is like. I'm not saying we shouldn't be a capitalist society, but like maybe we need to be a Capitalist slash Socialist society. You know. I don't know. (PIR 4, IDI). |
|  | There is existing support for initiatives that integrate healthier food options into existing services such as WIC benefits at farmers’ markets. | Community incentives and initiatives | so I think and I know there was initiatives, I don't know if they're still in place to like allow people to use Wick at like farmers markets and like fruits and vegetable gardens and even get like a rebate or get like money back. But I just think more initiatives like that where it's not just like low cost or ideally free, right for these healthier alternatives, but that there also is like an incentive like, oh man, I'm not just going to get this for free, I'm going to get this extra thing. Yeah. So I think just working with, you know, different kind of. But it's hard too, right? For like the food banks and for shelters and things like that because they're not operating on a really big budget. So that's, that's what's tricky. And that's where I think you need like the policy piece to support it. (HP, FG 2) |
|  | This includes the difficulties in balancing capitalist profit motives with public health needs. It also highlights the difficulties faced by food banks, shelters, and other community services due to tight budgets | Systemic challenges such as the capitalist system constraints. | o I think that this has to start not just in a household level, but I think, like the State, and the Government has to be way more committed to the the health of our people, and obviously because they are there are already like contingencies and contracts in place is gonna be extremely hard for, like us to figure out like, whose responsibility is it. Because the government is going to say, Well, that's the company's responsibility and then the company is gonna say, well, if the government ain't gonna put no sanctions on us we gonna do, we wanna do. And then the company is gonna say, well, the government allowed it. So it's gonna always be these checks and balances that these dances that's gonna happen. But I do think that the companies that are providing us the goods, and the government should be actually educating us on this. But because it's a capitalist society, it's hard to do that like we're not a Socialist society. So it's kinda like asking a capitalist, a capitalist society to care about people in order for them to lose money .. which is gonna ultimately cause them to lose money. That's really what it is. So II think the First Level is like. I'm not saying we shouldn't be a capitalist society, but like maybe we need to be a Capitalist slash Socialist society. You know. I don't know. (PIR 4, IDI). |
|  | This theme highlights the need for more educational and structural initiatives | Need for educational and structural changes. | but I don't think other than Michelle Obama changing the whole menu at the schools, or whatever when she was the first lady that wasn't really talked about or expressed, or, you know,… (GBO, FG 1) |
| Financing | This theme emphasizes the importance of initiatives that go beyond offering low-cost or free access to healthier food options by also providing incentives such as rebates or additional benefits. | Incentivizing healthier food options | So I think and I know there was initiatives, I don't know if they're still in place to like allow people to use Wick at like farmers markets and like fruits and vegetable gardens and even get like a rebate or get like money back. But I just think more initiatives like that where it's not just like low cost or ideally free, right for these healthier alternatives, but that there also is like an incentive like, oh man, I'm not just going to get this for free, I'm going to get this extra thing. Yeah. So I think just working with, you know, different kind of. But it's hard too, right? (HP, FG 2) |
| External Pressure | This theme highlights the disconnect between business profit-driven motives and consumer health. | Commercial interests vs. consumer health | So respecting those great points that everybody is talking about. But ultimately, I think we feel to realize that those that are in business cannot. could not care less about what's on these packages or whatnot. Their end goal is to make money (PIR, FG) |
| Inner Setting |  |  |  |
| Access to knowledge and information | This theme focuses on participants’ knowledge on the amount of salt/sodium in food. | Lack of knowledge on the amount of salt/sodium in food | But a lot of people don't know that you get more salt from the barbecue sauce than all your sauces. So it's like you really have to be careful. What's it? Say, the normal amount that you put in your salad is like 2 tablespoons, I think. Gosh, so your salad needs to be smaller if you like it more wet, you know, with the dressing. (RMA 5, IDI)    Salt is in everything if you're eating bread, salt in it - You can be drinking your coffee, and your creamer has salt in it - like you actually don't know what has salt in it. Or the sodium content of it. And I don't think that we. as African American consumers, actually look at that part of it. It's more of just the salt that you add to your food, not the salt that's already inside of the food before you actually prepare it. Even our seasonings it could be. It doesn't have to be seasoning salt, but 9 times out of 10, it has some type of sodium content. (PIR1)  I don't think people realize [things] like lemon pepper seasoning [is] so like salt. Like I mean, like we'll grab certain ingredients not realizing all of those ingredients have salt in it versus say, going for like the powders, and then having maybe, like one base salt. There's like a somewhat of the conditioning of just believing that certain things just don't have salt like. (PIR_FG1) |
|  | This theme discusses participants’ limited knowledge on the association between sodium and physical health such as gout, hypertension, and heart attacks | Lack of knowledge on the association between sodium and physical health | so [focus on] causes say like if it was heart attacks, you know what I mean, or something like that. I think we have to, the way you have to probably put up posters. Probably every medical clinic has some. You know salt is not good for you. We have to get like the same kind of way that they did with smoking cigarettes until they just had to go up on the prices, you know. Warning. Salt. you know, can cause kidney failure. It can. You know I mean the things along that I think we need to be more aware of the cause of them, because you know what the bigger ones are doing is showing the little ones how to be consumed with salt all the time. And that's not healthy for them. (RMA2)  If you started some sort of campaign that just share the knowledge of the hazards [of consuming salt], you know. How it causes hypertension. You know what I mean. If you just steady - kind of beat it home. Salt causes hypertension. Salt causes hypertension. (RMA2) |
|  | This theme describes participants’ call for better education and training for sodium management and the challenges associated with high cost of health and nutrition classes. | Access to available education and training | most people do have their smartphones and can access online information. So is there an educational video? (HP, FG1)  And we do have care coordinators also who can help with coordinating care and educating them on those things. (HP, IDI2)  I've learned a lot more in my .. going to the doctor more (RMA1)  like the gym nutrition classes and stuff like that. It's expensive, which also leads back to my first thought of … I think it's sometimes the lack of knowledge or the lack of expressing stuff is intentional. Because why are [there] health[y] things? Why do we have to pay for that type of stuff. (PIR, FG1) |
|  |  | Clear need for education and training on sodium consumption | But it is education. In that sense they become aware of that. These are not just numbers, and this is something that can affect your life overall. So so educating is education is a key key aspect of oh, and and following up on that is also another key aspect before they have a stroke or counter type, or anything like that, you know. (HP, IDI2)  So sometimes information like you just said what they're eating, not necessarily what to eat, or even saying what not to eat, but letting people know the salt contents, salt or sodium of what they're eating is so high, because then they'll say, Well, I didn't put any salt on anything, but the stuff you ate was loaded with it. (RMA3)    I think the first activity is education we have to be educated. We have to be educated on the harms. I mean, we know, but we don't know. Once we know, we gotta start doing something about it. I think like in our church communities where we're there. (RMA1) |
|  |  | Faith-based education | I just find it interesting how bread is used a lot in the Bible. And I think if your church communities will explain a little bit more in regards to even the intake of what we're living on every day, I think [it could] be a teaching point and how to live abundantly. It's not just about money that I think the church can help. even when we have church events. Some churches do like Keep some people grow some potatoes in a garbage. Can you grow mint leaves like you can make so many things at home. And he showing his video to people. (PIR1)  So maybe thinking about what training needs to be done in congregate settings, for the people who cook in congregate settings, and how do you reach into the congregate settings to do that training. (HP, FG1) |
|  |  | Food labels education | the most interesting thing is you don't even see on the label anything about salt, you see, sodium sugars and things like that. So what I would like to see in the community is really, the breakdown of the label of what's actually in our food will be helpful. (PIR, FG1) |
|  |  | Lack of knowledge in the community | I don't know what I'm doing wrong or what I'm not. (PIR8)  I don't know if exercise helps. Or I don't know if sweating helps I don't know if, like increasing your water or hydration helps. I don't know for sure. But I am curious to know if either of those things that I mentioned help. (PIR7) |
| Available resources | Resources are available to implement and deliver the innovation (funding, space, materials & equipment) | Health care options | I don't see those type of things. The only thing I could say is people are attending they're checkups that's, I think that's the only way I see that they're getting it or being more involved with the programs that they have with their health insurance, and therefore those that don't have health insurance? That's questionable, you know…Oh, well, for older people pretty much, most of probably going to the doctor now, because of all the different things that develop with probably with salt intake. (RMA5)  They do that (in) tapestry already, which used to be Heartland. You know, they have providers and I think a case manager or something where they actually set up once a month and they teach them how to do healthy cooking. So it's just someone in the clinics who would be able to do it. (HP, FG1) |
|  |  | Lack of healthy, low-sodium food sources (e.g., Grocery stores) | I would say whole foods. But you know they just recently moved the whole foods that was off of 6 63rd . They shut it down and a lot of people I mean, the community is upset. When whole foods went to that part of Inglewood it was a big thing, because whole foods is coming to the south side, and then whole foods is going to Inglewood like what? That's how we knew that the neighborhood, the Inglewood community, I don't live in Inglewood, but the Inglewood community was on the brink of change. But now that whole foods is gone? They were trying. (PIR6) |
|  |  | Restaurants | There are certain places, I think, more restaurants now, the newer restaurants that open up and style. They don't cook with salt. They don't cook - remove seasonings. They allow you to season your food to taste, and I think that's the way it should be, and you know what? Because they were doing it. It started having me do it, too. So that's a good thing. (RMA2)  Yeah, we have, you know we have more stores coming up, that are more on the healthy side, more organic. farm raised food, no pesticides, and all this stuff, so we can go to those types of marketplace. There are more restaurants that have like a Vegan restaurant, you know. (RMA1) |
|  | Funding is available to implement and deliver the innovation | Funding | *(only quotes from HCPs)*  Everything that happens needs to be funded through a grant or something because, you know, the clinics are barely trying to keep their doors open. And, you know, they have a lot of patients who are self-pay who really don't get paid for their thing. So, you know, we would have to focus on or you guys would have to focus on finding grants for all of these extra stuff you guys would like to add. (HP, FG1)  More funding, because, remember, a lot of the healthier things are a little bit more pricier. And at the same time, too, you do need non perishables, because that's the reality. You know. So it's just really the funding cause, you know. (HP, FG3) |
|  | Supplies are available to implement and deliver the innovation | Materials & Equipment | We can print out dash diet information sheets from the medical record or from just going online to print it out. But we don't have a lot of other information. (HP, FG1) |
| Communications | There are high quality formal and informal information sharing practices within and across Inner Setting boundaries (e.g., structural, professional) | RMAs as a conduit for information | I'm part of the Health Ministry. Sky high, their blood pressure sky high, be liking it okay, what did you eat today? And when they say, oh, I had a bag of chips, and then I had a pickle. and I had a can of pop. I'm like, okay, look, look at all this salt. You had a bag of chips a can ofa pop and a pickle? You had a whole lot of sodium right there that you wasn't necessarily aware of but it was sodium intake. (RMA3)  It's almost the same thing I feel like I've heard about when you wanna work out like getting a workout, but be like just not being in something specifically new or not being alone is very, very helpful, like you have the teamwork aspect, and you have somebody to check you… (PIR, FG1) |
| Compatibility | The innovation fits with workflows, systems, and processes | Many small ways to fit in discussions about sodium consumption in a health care visit (but always fighting limited time) | I think the conversations should start before the provider enters the room. And and that's really looking deeper into like social determinants of health that's usually acts by. You know the care team, whether it's the nurse, the medical assistant, the Dietitian case manager. So I think those conversations can start prior, and having at least an understanding. So when the provider goes into the room, being a provider, would know, you know what's the greatest, what are the greatest barriers or where to kind of focus? You know, provide kind of focus, support. So that's just for me, I think that really, providing that appropriate work flow because a lot of times you can ask the questions. (HP, FG3)  I was champion for the hypertension control in this clinic. So what I do when those patients come is first thing is of course we check the blood pressure. and second thing is, we make them wait these 10 min and recheck the to make sure it's the right reading. Third thing I do is I do it. A beautiful handout on dash diet. If you are familiar with that and dash diet details I give to them. (HP, IDI2) |
| Culture-  There are shared values, beliefs, and norms across the Inner Setting | This theme focuses on the challenge of reconciling spiritual symbolisim and physical health impact of salt | Salt in faith-based culture has mixed meaning | I agree with FG9 cause as I was thinking about it, even just the 2 references that we brought up talking about like being the salt of the earth, but also how Lots wife was turned to a pillar of salt? It's like the connotations are different, like nobody wants to be in the context of, you know, being turned into a pillar of it like, clearly you do something wrong as opposed to, you know, being the salt like, you know, that's something that you want, or that's something that you know is seen as in a positive light. So to FG9 his point. .. I believe Salt has a place, you know, in the faith-based community. It's just going to depend on the context, on the environment, on the usage. And then I like the word that FG9 used that balance sort of thing. I think that that's applicable. (PIR, FG1) |
|  |  | Role of food in faith-based culture | So I know in faith-based communities, they're like a lot of meals. They're like conferences and things of that nature. So food is always involved in community spaces. And it's good food generally. So like, there's a lot of salt that's used for that. (PIR, FG1)  I'm just going off of history, and with like after church dinners like gatherings from the community sense. Even just family gatherings. Wow! Just the the rich history of like passing down recipes passing down just like similar, like. There's like a standard with certain dishes. With that, a certain standard with taste, and stand with that the standard with taste, the standard with salt intake. Thats a reason why, like a lot of the stuff just ends up getting perpetuated. (PIR, FG1) |
|  |  | Cultural traditions and resistance to change | So I know in faith-based communities, they're like a lot of meals. They're like conferences and things of that nature. So food is always involved in community spaces. And it's good food generally. So like, there's a lot of salt that's used for that.  I'm just going off of history, and with like after church dinners like gatherings from the community sense. Even just family gatherings. Wow! Just the the rich history of like passing down recipes passing down just like similar, like. There's like a standard with certain dishes. With that, a certain standard with taste, and stand with that the standard with taste, the standard with salt intake. Thats a reason why, like a lot of the stuff just ends up getting perpetuated. Because if say, you're the cousin, and you decided to not put any salt in the Mac and cheese, because you knew there was already cheap, like salt in the cheese. But like it's, the actual thing … You're scrutinized for going against the grain for like helping the people get better. so I see it more like in that colloquial, like familiar community, like cultivating, and like that, traditional like recipes, traditional, a traditional flavoring of things having to be salty, or even if has not even being referred to salty. Just good, like good means, salty. So that's the point you have to kind of cut against that and cutting against like those traditional things [is hard] , because for most people that is at the forefront of just like they want a code they want to continuously pass down. Well, this is how my mom taught me this. She learned it from my grandmother, her great grandmother, and I'm not about to stop that.(PIR, FG 1) |
| Mission alignment | Evolving health focus to cover broader health initiatives including health literacy carrierd out by Pastors4PCOR, TRCDO, and Pastors. | Evolving comprehensive health intiative within the community | because like P4P, and like TRCDO offers a lot of like health literacy classes and opportunities for engagement. They just had [somthing] like Man Church, where, like people came out, talked about like prostate cancer, and all of us like health stuff, mental health, and all this other stuff. And then they did the same thing for the women. And we try to like meet the needs. (RMA 4, IDI).  you know, as far as P4P or my like, my church it could be both, it could be like around TRCDO or your church. I think we can definitely like pull it together. We have, like a health and fitness ministry. The leader over there is actually like a personal trainer. He owns his own business. He actually is training a lot of people at church. So I think if we can, we get a lot of people on board like a campaign and make it a big thing. I think it will be you know, we will make it fun. I think if we can like be real with people, I don't think it has to be like a bunch of like pandering to people like, oh, I think we should be like real and like, let's talk about. Let's think about it like, let's let people ask questions. So I think it can be done in a different manner. Manner if we all like, come together and like brainstorm, how to work it. I think it could definitely like be a thing. Our pastor is big on health as well. So he'll be like. No, we still need to be like, he don't eat a lot of stuff, but he'd be like, no, that's not healthy. (RMA 4, IDI). |
| Structural characteristics | Layout and configuration of space and other tangible material features  support functional performance of the Inner Setting | physical infrastructure | And then you also have factors of like, food deserts and like safety and transportation. So like your fresh food is far, you have to get it from A to B, not only is that fresh food more expensive, it's more expensive to go get it, and then you might not feel so safe doing that. (HP, FG1)  Yeah. I was gonna say something similar. I will add, I agree. And I will add to like healthy, especially in the food area, like healthier foods generally are more expensive. I grew up in a low income community. So we had a very health, conscious family, so to his point, like the healthier, like options with like organic food and all that weren't in the neighborhood. I grew up. We had to drive like 20 min, so the suburbs to get to a whole foods are Mariano's or the cheaper version of that, because we would have loved to be shopped at a Mariano but we couldn't afford that. So we had to do our best with, like the all these Jewel Oscos, you know, and just kind of pick and choose what we could find from there. Just because the resources weren't available like we had the knowledge, but there were no resources. But I agree with everything, FG7 said, and I just wanted to add that to bit. (PIR, FG1) |
|  |  | Work infrastructure | And then community health workers, you know as well, um, I know we have an initiative specifically we got funding from Hrsa, those community health worker training program, and we did a very conscious effort of recruiting trainees from our community. Um, so it could be, you know, community health workers too. So I think like a combination of having, you know, health care workers that are licensed to do X, Y and Z, but also people who are non health care workers or who just are involved. And as a community health worker, for example, I guess that is a health care worker, but that focus is more on the community education, um, but folks from the community and trusted leaders. And um, I don't know, I feel like I'm rambling, but those know yeah. (HP, FG2). |
| Relative priority |  | Comorbid health conditions | Then other, usually people have many other disorders that we're trying to treat at the exact same time. And also like priority, to be completely honest. Like again, if there's like depression or substance use disorder, like I have to go with what's, you know, going to, I guess mostly like kill the patient first, it's not the best way to put it. But sometimes that's also, not every visit, but sometimes that is what I have to like prioritize. Even though I get it, diet is really, really important. It's just, we've got a lot of things to cover in a really short amount of time. (HP, FG1)  And a lot of times patients…get to the provider and they want to bring a whole other list, so it's like they really don't have time because what they're really coming in for, they can't even focus on that because they want to tell the provider everything else that they need. (HP, FG1) |
|  |  | Other nutrition concerns | And to be honest [I think] there is more [to] sugar….Most people don't look past that very first line [calories], and it's even strategic, and how they even set it up like labels, cause you'll have the calories in big old [font]. But then, when you get down to sodium and all these different preservatives its all in very fine bent lines, so you can't. (PIR, FG1) |
| Individuals |  |  |  |
| Capability | This focuses on recipients’ knowledge, skill and confidence in managing salt consumption and reading nutrition label. | Recipients | I think I would say, I don't like really consider it when I'm eating out. So whatever the food is made with like, that's you know what I'm eating. But when it comes to cooking I don't use like regular salt I use like sea salt and like maybe some seasoning. But I tried to not use as much salt when I cook. So that is like the only way that I intentionally try to measure it (PIR 2, IDI).  Salt intake cannot be controlled. I disagree with that. I disagree with that, especially if you cooking . If you, if you preparing your own meals right, we control what we put in preparing those meals so yeah, it can be controlled if we prepare our own meals. Right? Now, we eat out, you know, not so much. You know we don't. We can't control that. You know, we out but we definitely can control what we are prepared (PIR 3, IDI)  Eat less processed foods as much as I can. Eating at home. As I said , you go out , there are those special cases, and I love to go out to dinner like everybody else, but you have to do everything in moderation. You know too much of anything makes you an addict. So I don't wanna be addicted to eating so many foods that have high sodium content are just not good for me in the long run (RMA 1, IDI)  I don't know a lot about it, which is why I I tried to cut it out I have one box of salt salt in my house right now that I've had for over a year. and there's still some salt in there like the box is not totally gone. But I've had this salt box, I wanna say, for a good year and a half, because I can count the times I had just a pinch. It can be too much . And so if I add salt it is just a pinch. I'm telling you I've taken it a step further. I don't even put salt in my eggs. I don't put it in grits, you know, when I'm making stuff that normally requires salt. I don't use it. I just eat it plain. Or I have to use something else, garlic, powder, or maybe onion powder to substitute (RMA 2, IDI)  Because I don't think people read labels. it wasn't until like maybe 2020, where I started to really like point in on how much I was actually taking, because I was working on the ambulance full time, you know, and I literally saw the progression from like working a regular shift to working in like a pandemic. So we were grabbing the first thing I could like to start with. … [and] at how much salt is actually in a bag of chips you'd be like, oh, like, you know, and like even a bag of Lays, [and] I had to like, train myself to relook. (RMA 4, IDI)  I'll go back to education a bit. If it says something else, I'll just automatically stay away from it. But I know now I need to still be educated on how to read these labels. What do they mean? But right now, if I see something like this, I'll just stay away from, because in my mind I don't need to eat that. I don't need to drink that because of the sodium. (PIR 3)  And we we don't know how to read those full labels on the back. If it's say 10 calories for 10 chips, that's what we looking at. (PIR, FGD) |
|  | This focuses on innovation deliverers (RMAs and health professionals) ability to educate and influence people’s dietary behaviors | Innovation deliverers | Yeah, I think I would. Personally, I feel like I am very convincing, and I think that I'm very like natural. I show up. I show up as myself everywhere like I showed up as myself today, and you know I'm I'm I really should be a comedian . But I think I'll do very well at explaining why it's important or how to useit especially if it's like user-friendly. My, personal view is a lot of senior citizens may like it but so long as its not if it's not gonna be like, you gotta do a whole bunch of steps. They're already out. But I think if I learn how to work here and like am confident in using it and can I kind of test that myself, I definitely would be like, alright. Y'all like this in the way like you know. (RMA 4)  Like I don't think I'm proficient, I kind of have a few good tips that I try to tell patients. (HP, FGD 1) |
|  |  | Recipients | I would. Oh. It would make me think about how can I cut salt. Well, it made me think about the other thing, and the reason why I have just not read labels as much is because I think for me it was very tedious, a lot for me to kind of read labels and things like that. So what I've tried to do is just make an overall lifestyle change. And so this, eating more fruits and vegetables and things like that, less processed foods, less fast food in general. My thought and hope is as I work more of those in [is] somehow some way it'll just be better for me as opposed to eating a lot of fast food or fried food or junk food or anything like that. Yeah, I think reading, because reading in general, or, yeah, trying to pitch how much was it? Every pack was just a lot, and it got tedious. And I was just like, let me just make overall change instead of trying to read everything, every single thing (PIR 8)  So it depends - what you're talking about, because I love salt. I absolutely love it. If we're talking about food. I love salt I can't get enough of. So , even if something is too salty generally, it's pretty good. So I'm like, Okay, I need to add a little salt to it, or something that has salt or a heavy sodium content. I love pizza heavy and sodium. Everything that's heavy in sodium. I like it. (PIR 1)  But there is this sense of. Hmm! Hold on! There is this sense of kind of sacrificing or like they're just already changing. They're already not doing heroin, or they're already, you know, like not doing cocaine, and I'm telling them to stop smoking, and you know they have very little as far as like pleasure kind of thing. So when I start telling them to eat more like a bunny rabbit. That is like sometimes a bridge too far, just even conceptually so with education. I do tell them there are factors that you can change that are in your control and their factors. You can't when it comes to like blood pressure. And all this stuff. And yeah, like, dietary choices are one of them. But I just feel like when I start talking about that. It's just unreal it like they. They just turn off, you know, when I start talking about like you, you really should decrease, you know, processed foods and all that stuff. (HP 1). |
| Motivation | This includes challenges faced by participants including (the inconvenience of dietary changes, effort required to understanding nutrition information, and preference for high-sodium foods) and success with gradual changes, sustainable habits, and patience in managing sodium intake | Recipients | I agree. And why I agree is because I believe that you know, just throughout our daily lives we are eating and consuming and not really paying attention about the you know how much of what ingredient - any ingredient - is in anything that we eat. So I do believe that some people , most people, are not aware of how much salt they could potentially be consuming. (PIR 6)  Right? Right? So I would have to say. I agree. Nobody really measures what we're doing if you have salt, you just putting it on all your food directly. Nobody, nobody measures that (RMA 1)  Okay. What helps me is, I like smoothies, and like meal replacement shapes and stuff like that. So that helps me as well, because [I’ll pu] out, though, like vegetables and fruits in in a blender and blend that up. So that helps me minimize my salt intake and also using other like salt substitutes.(PIR 3)  but I'm thinking I'm optimistic, and I'm grateful to be learning this information now, so that in my mind I can make the necessary changes, and I can discipline myself, although life can be happening like. I want to live long. You know what I mean. Life may be lifelong. I don't want to have what my dad has, you know, so I have to take the discipline, and I have to take the responsibility to get the information, receive it and go from there. (PIR, FGD) |
|  | Innovation deliverers identified challenges like variability in individual responses to salt and lack of control over the effects of salt on blood pressure. They also emphasized empathy, personal experience, and the potential acceptance of salt substitutes through taste tests and food preparation demonstrations as effective strategies. | Innovation deliverers | But we don't have a lot of other information. And I have to say for myself, this really is a barrier is because not everybody responds strongly to salt, like the link between high salt and high blood pressure, it's very true for some people and not that much true for a lot of other people. And if I feel like people don't have that much control over it, I don't emphasize it too much, just the reality of my own practice right now. But if someone does have persistently high blood pressure despite the meds, I will say try to avoid salt, here are some, you know, ways to try to avoid salt. But it's not one of the first things I go to. (HP, FGD 1)  I think that if they tasted it, like if there was a taste test and they could see that it tastes pretty much the same. Like, I mean I use salt substitutes, so I'm biased because I think it tastes the same. So I always push people to use it, but I think that they're scared of it kind of because they're not used to it. And it goes back again to having them taste the food, show them how to prepare the food, give them alternatives. But, um, I think it's possible. I'm not really too sure. (HP, FGD 2). |
| Opportunity |  | Committed decision making and meal preparation | But I think if people are committed, even if they have to be stricken over the top. I think it's possible. I just don't know if folks are willing to be that committed to doing that type of detailed work on a daily basis to do it, but I do think that you can measure your salt intake (PIR, 7) |
|  |  | Use of technology | …Not most few of them do sign up for those, and so there, some of them are technically savvy for those people. I think it will be very effective. (HP 2) |
|  |  | Inconvenience of reading labels | I mean, when I see nutrition facts on stuff that I purchased from the grocery store. I'll look at it. It doesn't necessarily impact if I'm gonna eat it or not cause at that point if I've bought it like, purchased it and brought it home I'm eating it or drinking it. And I'll be honest if this is on something that I like buy out - like fast food I'm not even looking at it. (PIR 7) |
|  |  | Difficulty of making informed choices when hungry | A lot of our patients, they just want to get some food in their stomach. You know, they're so hungry that they don't even really, that's so secondary that it becomes difficult. If you're really hungry, you're going to eat whatever is available, you're not going to even think about like, oh, is this high sodium? Like, maybe I should put it down. You know, you're just hungry, you're going to eat what you can. (HP, FGD 2) |
|  |  | Financial challenges and limited access to healthy foods | But I get it. Sorry. I just thought about something this morning. My niece, I have a niece who's like 2, and I also have a goddaughter who's 3 now, my niece. I remember I just saw a picture of her eating like chips or something like that. But then I was facetiming, my goddaughter and she was eating like blueberries or something. And I'm just like, Why is that the case, or whatever? But its money issues - like it's cheaper to get the best stuff. And so if that's what they can afford, then they just don't get it. And so no, that's [not] good. Should be the best. (PIR 8)  and I think it's access and it's finances. You know, a lot of our patients, even if they had the finances, they still would be going to fast food a lot and I don't think they know, but the two of them together makes it much easier to access, you know, highly processed foods with a lot of sodium. (HP, FGD 1) |
|  |  | Challenges with technology (SaltSwitch) | It may work, but couple of challenges that we have is, you know, most of the people are not very well familiar with the smartphone. Even now, you know, they have the basic phone, but not the smartphone or the apps. Many of them are not, especially, of course, the older age group that still has challenges, you know. So but for other people, many of them have our own portal and everything, so most of them do sign up. Not most few of them do sign up for those, and so there, some of them are technically savvy for those people. I think it will be very effective. But for people who have a bad connection, you know, even when I try to do a telehealth visit with patients through the video to the zoom, and many people are still not first of all, comfortable with it. Secondly, they're not. They don't have the best Wi-fi connection available, and thirdly, they're not comfortable using it. So, I would say, maybe 20 or 30% of the people with hypertension may benefit from that. (HP 2) |
| Need | Awareness of the impact of salt on health issues such as high blood pressure, heart problems, gout, and obesity, often triggered by personal experiences like developing hypertension after childbirth. | Awareness of health risks associated with salt intake | When I had my first born, when I was 21, I had hypertension after she was born. And that's from eating all you know, going out and everything. Its not just the weight factor, it is just the intake of it, being in your system and it depends on the individual (RMA 5, IDI)  One of the consequences - are we just passing it on from generation to generation? If nothing is changing, we're not implementing anything (PIR 3, IDI)  Long-term is really a myriad of things like ... I don't know the extent of the ailments and the diseases that plague my community, but I know that it's a lot of 'em and I know that a lot of them have to do with not having healthy options (PIR 6, IDI) |
|  |  | Knowledge gain from observing the effect of salt | I do not. I do not use regular salt based you know. Regular salt, I would say, like Morton salt. I do have the Himalayan salt but I use that sparingly, and I would say, probably once in a blue moon, maybe one to 2 months, I'll use it. so I just pretty much think about how to use it. If I use any salt, it's pretty much. It's pretty much like Creole seasoning know. Cumin powder, you know those type of things. I never buy anything that says, you know, salt, you know, like with the different seasonings. But it's decision making. I'm not gonna use it because I think I prefer to growing up. You know, in my family we have strokes and high blood pressure, you know. I don't want to experience that. So that has been controlled, as you know, knowing myself and my body and my health you know you know what you can and cannot eat that may trigger certain things in your body. If that's gonna be harmful. (RMA 5) |
|  |  | Convenience and availability of fast foods | The fact that it's prepared and it's called fast food for a reason. So you can grab what you need to grab and go, but not knowing what they're putting in that food. (PIR, FGD) |
|  |  | Lack of access to healthy foods | So we were grabbing the first thing I could like to start with. … [and] at how much salt is actually in a bag of chips you'd be like, oh, like, you know, and like even a bag of Lays, [and] I had to like, train myself to relook. That's why I feel like crap like I literally feel like my heart beating through my chest. And I was like, I need to go to the doctor. And she was like, Yeah, your blood pressure is really high. And I'm like, Yeah I can literally feel my heart like skipping a beat. And she's like, What are you eating? And I'm like, no, we just we stop. at Mariano to grab luncheon, you know … (RMA 4) |
| Implementation Process |  |  |  |
| Adapting | Modify the innovation and/or the Inner Setting for optimal fit and integration into work processes | Salt reduction is not going to help everyone control their BP | And yet for those one in four, it makes a huge difference. So I say try to cut it down, here's ways to cut it down, see if it makes a difference. (CHC, FG) |
|  |  | Community members are not always comfortable in the health care setting | …a lot of times a church or a community setting, that's what we feel most comfortable. We really don't like going to the doctor. It's not good sneezing, but somewhere where you go every Sunday or one day a week for Bible study, or a choir on a Tuesday night - you have somebody there where you already comfortable with this setting. (RMA1) |
|  |  | The presentation of information (i.e., in the form of 5 mg) is not digestible to people | But that 5 5 grams a day, I think even though, like I'm not gonna measure I think. but hearing that will make me - like will make me adjust. And also, like as I tell people like, Hey, it's as simple as when we have the salt Shakers don't shake so many times. You maybe just shake once or twice, and that's the practicalness of that's that's translating it in a more practical sense than say 5 grams like a teaspoon cause no one, no one's gonna really care too much about that… (PIR, FG)  I would say maybe advertising this stuff in those seasoning aisles in stores. It's one of those things you can put right if you are really serious about it, and it is such an urgent thing you should place these things in the marketed areas. You should have something about high blood pressure, sodium, intake, sugar intake in these aisles? Just so that people are aware of it because if [the info] it's only available via Google or via resources in which you have the person they look like It's not going to really be sought after versus saying, Hey, there's a little thing right next to the pricing label talking about this amount of sodium. Be wary. Be wary of this. If you experience a high high blood pressure, we suggest maybe the low sodium products. (PIR, FG) |
| Doing | Implement in small steps, tests, or cycles of change to trial and cumulatively  optimize delivery of the innovation | Momentum is needed | Second step is, once you got the education you have to, you have to. You have to do something about it. We have to study it, practice it, and then we teach it to other individuals, then it gets in your DNA, and it becomes a habit, and once it becomes a habit, it becomes a lifestyle. And then that's when we can change and be healthier. (RMA1, IDI)  if they're more active. Think that they'll wanna use less salt, because they'll see the benefits of being active, of moving around versus you, checking all this out, and you don't feel good, your head hurt, and all that, and you don't want to do nothing (RMA3, IDI)  But there are different things that we eat that turn into sugar in our bodies, and that's what our body is craving. So I believe it's the same thing as it relates to salt. That there are certain foods that we eat when you, for for instance, me and my wife, we went on like a Vegan diet right? And then we came off the weekend. I think we did it for like a month we came off of it, and it was like everything tasted too, too salty like everything was just like.. it was almost like we our palettes were cleansed and we were seasoning stuff like how we always done it. And it was kinda like, yo, this is way, too much salt, like I actually can't eat this. I can't take this. And so I do think that once a palette is clean once you start to eat different foods it's almost like your body trains itself or recalibrates itself and shows you what's not normal (PIR, FG?) |
| Engaging (Recipients) | Attract and encourage recipients to serve on the implementation team  and/or participate in the innovation | Providing education and activities | maybe like more awareness events. Something that maybe just not strictly informative, but also has, like a aspect to like a family play, you know, activity, part. I think that would be great. (PIR, FG)  So I believe, like the education portion is definitely [value] seminars and things like that money focus, or like. Even, I think I saw some stuff on how to grow your own garden, or something like that, but I think getting that knowledge and that sort of way will be good for free, though, like free, you know, seminars. (PIR, FG)  Oh, oh. oh, maybe if they had like like cooking classes to show people how to like, actually prepare food that will still be flavorful without using as much salt that will probably be helpful (PIR, FG)  Activities. More health fairs, you know. I know we went virtual, everything online, I think, having more health fairs, educating the community especially in the black community and then give .. and you know, educating and also providing resources, maybe giving people blood pressure cuffs, you know. Yeah. (PIR, FG)  Live visuals. They're somewhat hit or miss, but I do think when you're seeing, I think there was once where I saw like -they did, like an actual demonstration of like, hey? They told somebody like, give me the ingredients like for, or give me a recipe for something, and I'm gonna show you like how much of everything [is in it] so you can see, like how much salt, how much sugar and then that person changed the complete recipe, because. like he had both like bowls full of, like a lot of different seasons. Because half the time we're going into seasonings flying where you kind of just like we just go off of just like instinct and muscle memory versus how much to actually, how much you actually need on something.  I feel most people in situations like this are more prone to being visual learners, and you don't know it's a lot until you actually see it. So you see that you're filling up like a big cup like this? With a whole bunch of sugar, and like a small thing like a small little like cookie like that's a lot of sugar. But you don't know, because all you see is the final product of the cookie? So it's just making people more visually aware of what is actually like being put in their bodies. (PIR, FG) |
|  |  | Engaging one-on-one, breaking down individual barriers | I think, dedicated diet…trying to identify patient, specific barriers right?...actually sitting down with the person in the room and saying, like, you know, this is what we're talking through, as far as best case scenario like. Is this possible? If not like, what do you think makes this difficult for you and then trying to work through them on on the specific situation? (CHC, FG)  And it's just a change of people, my state 1 one person at a time, you know. You know it's a numbers game, you know. You talk to 11 that believe you talk to 20 and you get one. You talk to 100 and you get 10. It's a numbers game. We just gotta get the information out. (PIR, FG) |
|  |  | Broad marketing | If budget wasn't, I think we definitely need like huge marketing. So it's just not like the people in my community. I think people to people in the community where my church is located should be invited to this = something like billboard like, let's, do you know? Like, let's do something, or like (PIR, FG) |
|  |  | Use opinion leaders | I don't think it should necessarily be like an RMA or somebody. I feel like we should be supportive. But somebody who like has some credentials like, you know people with the letters behind the name like I got my own letters behind my name now, but like somebody who's well versed, and who can like speak to all demographic and who could be real like when it's on the patient education. (RMA, IDI) |
|  |  | Start early | And eating habits are formed young. They are really difficult to change when you're older, really difficult. So if you grow up on your healthy snacks, more likely you're going to crave those later. But if you just get diagnosed with X, Y, Z, and then you have to change all your eating habits, that's a big ask. Like that's really, people can do it, but that's a bigger ask. Get it in the schools, we're already having to provide food for schools, so do it. And I say make it like whatever, like the gardens, all that good stuff. There's so many schools that do that now. (CHC, FG)  Or your local boys girls club things of that nature and then impact them. Educating the kids, too. First and foremost, get out there…They gotta do activities to get their heart pumping. It's a move, you know. (RMA1, IDI) |
| Engaging (Deliverers) | Attract and encourage deliverers to serve on the implementation team and/or to deliver the innovation | Get researchers/people with particular expertise involved | I think I'm speaking with him now ! I could see yourself and others leading other people to come in and get the knowledge where they could share the knowledge and be representative, to go out to the community to share that particular knowledge. (RMA5, IDI)  I think you would need a counselor. You need someone. When I say a counselor [I mean] because people need to come in and talk about it . It's a mental thing of why you eat what you eat , why you eat much of it. Oh, you know what I'm saying when you come to salt, how much intake do you think you eat daily? So you need those types of questions to me [via] to counselling. I would say, someone like a behavior specialist. (RMA5, IDI) |
|  |  | Use opinion leaders | First and foremost, the pastor, the head, of the household, the head of the church, you know it has to come from him [then] through the other leaders, through the rest. Once he’s vocalized it and spearheaded. Because let's be real a lot of times, you know it doesn't…it has to be from the top down, going down, and sometimes the bottom up is good, too, because that's that's young minds coming up talking about it. But in the beginning it has to be coming from the forefront. You talk about a church community, or a coach talking to his players, or whatever. But it has to be the leader. (RMA1, IDI) |
| Reflecting and Evaluating | Collect and discuss quantitative and qualitative information about the success  of implementation and/or the innovation |  | One of my favorite things is when I see people using a lot of salt. I tell them. You know, if you drop dead, Morton salt is not going to stop selling that one box. and sometimes they'll get mad with me. But they get the point. They ain't gonna miss you, 'cause there's gonna be some more people doing it. So you may as well take care of yourself. (RMA3, IDI) |
| Planning | Identify roles and responsibilities, outline specific steps and milestones, and  define goals and measures for implementation success in advance | Start with providing knowledge | I think the first activity is education we have to be educated. We have to be educated on the harms. I mean, we know, but we don't know. Once we know, we gotta start doing something about it. I think like in our church communities where we're there. And you know, it used to be, you know when I was a kid at the church, you know somebody's downstairs, France something, and you had dinner and all that is gathering is beautiful, and it's great. Now let's have a health ministry. Let's have a walking ministry. Let's go out. If you go out to evangelize, you know. Let's get these steps in. Let's walk a little further. Just 2 blocks around the church. Let's go. Let's go 6 blocks down and come around and and and knock out 3 miles, or whatever we can do. So I think the first step is education.(RMA1, IDI)  But I would definitely say to start off with plain research. Really doing a deep dive into what we're eating while we're eating it? Where is it coming from? Who is making it? What is the pro? What is their process and making it and researching like where their actual labs are and where their farms are - even researching like , if these companies have been sued right - like have they been sued for the fact that you know, for Salmonella, or all of these things, just to see what comes with the food like like, what? What are they spraying on [it]? A product cause all of these things also - can attribute to these chemicals that are in our bodies. So yeah, I think to start off with the first. The main thing we need to do is like, have communities of research. (PIR, FG) |
| Tailoring strategies | Choose and operationalize implementation strategies to address barriers,  leverage facilitators, and fit context | Investing across the lifespan | I think it needs to be implemented in the school in all levels. Not just your health class. When you learn about menstrual cycles. It needs to be .. what are your core classes like when you take algebra and things of that nature in the school? You should also take nutritional chemistry like, know what this and then at the college level, you're changing your mindset overall (PIR, FG) |
|  |  | Patient-led goal for change | Yeah, if you could get people to think goals. Sometimes maybe weight goes. Sometimes blood pressure goes or a goal to say, Okay, I'm gonna work out 3 days a week for 15 min. something like that, and then when you see them again, they'll be like, well, II made them 3 times and you know, and they'll feel proud of themselves and maybe want to continue (RMA3, IDI) |
|  |  | Focus on the positive aspect of health | And we just have to stress that we need to live and not just live, but live and flourish and be happy and be healthy. You know I don't just wanna live dormant. I wanna live positive. You know, I wanna be able to see my grandkids kids I wanna be that type of laser guidance. Going to [getting] us better, all better, all health (RMA1, IDI)  People being offended. Thinking you’re talking personally to them… We talking about eating too much fried foods and this that and the other Some people could be offended, couldn’t they? Don't wanna be told anything, but that is an individual issue with the message. (RMA1, IDI) |

Abbreviations: CFIR. Consolidated Framework for Implementation Research. COMBI-SR. Communication for Behavioral Impact for Sodium Reduction. FG. Focus Group. HP. Healthcare Professional. IDI. In-depth interview. PIR. Potential intervention recipient. RMA. Research Ministry Ambassador.
