## Supplemental File 4 for "Contextual factors and implementation strategies for a multi-level community-based sodium reduction intervention in Chicago’s South Side: A qualitative study"

| **Supplemental Table 3. Strategies for Implementing the COMBI-SR Program** | | |
| --- | --- | --- |
| **ERIC Domain and Construct** | **Definition** | **Illustrative quotes** |
| Conduct local need assessment | Collect and analyze data related to the need for the innovation | I think if you have buy-in from executive leadership, and the way you do that is by showing them the numbers. Like, I know our hypertension numbers are horrible; they're abysmal. Someone may not be—I don't want to be morbid—but it may not be tomorrow. But this is a big problem, and it's going to really negatively impact our patients. So, I think getting buy-in by showing them the impact is crucial because most people are there because they care about their patients. And a lot of our patients face many different challenges in their lives. So, something like that would be critical. (HP, FGD 2)  I think again, like, if you have the resources to understand, like to spend the time and understand, like what people specific barriers are, then you could target them (HP, FGD 3) |
| Tailor strategies | Tailor the implementation strategies to address barriers and leverage facilitators that were identified through earlier data collection | tailoring the plan through a team approach like with the nurse. And you know, maybe like a little breakout like kind of nutritionist in the office if there is one. So yeah, tailoring it, individualizing it (HP, IDI 1). |
| Promote adaptability | Identify the ways a clinical innovation can be tailored to meet local needs and clarify which elements of the innovation must be maintained to preserve fidelity | I think that's hard because sometimes it's just a lot of it is like geographical, so I think that that piece. And like again, I go to a bunch of shelters, right? So something that's like accessible to one group of people. Like I could do the whole city, so it's not reasonable for a different part (HP, FG 1).  But then again, they need to figure out if they're waiting for people who are going to have dramatic changes in salt intake before knowing whether or not they need to commit to that. So this is why it's a little bit harder. And yet for those one in four, it makes a huge difference. So I say try to cut it down, here's ways to cut it down, see if it makes a difference (HP, FG 1). |
| Identify and prepare champions | Identify and prepare individuals who dedicate themselves to supporting, marketing, and driving through an implementation, overcoming indifference or resistance that the intervention may provoke in an organization | For getting community leaders involved, there's going to have to be some type of probably incentive for them to do it as well. And I think repetition is key too, just like talking about it all the time, you know, bringing it up. Even if it's not the most important thing that they're there for that day…I don't know how important hypertension is when there's like shootings going on and all that stuff, you know? So I guess, like I said, again, repetition and community education would be probably the best things (HP, FG 2).  You have to interview them and make sure that they're the right fit for your program. Because if you're just trying to hire somebody just to get the survey done, then I don't know, it's not going to really work. Because, you know, patients need to know that you care for them so they can care for themselves sometimes. Because a lot of times, I have patients who have no family, who don't care, so they're like whatever, I'm going to die anyways, right?...So basically that's what you guys would need for the program, someone who has passion from the heart, you know? (HP, FG 1) |
| Build a coalition | Recruit and cultivate relationships with partners in the implementation effort | The South side has the highest net density, like more churches than any other community in the US per population. So going into the churches which are trusted spaces to work with pastors, to work with church kitchens, thinking about where are the places where people come together to cook together…The main distributors of food knowledge would be where communities come together to eat and figuring out where those spaces are.  I think the more we can…partner with the community and people who provide the food [such as Feeding America], um, and like you said, like food is medicine and that's really like taking off.  Work with CPS to make sure that there are salad bars in the schools like structurally, is each school able to serve low sodium food or there is one high school on the south side where they didn't have a school kitchen, it was a charter school. So they literally served cold cut sandwiches every day and they couldn't have a solid parks, there was no place to have it. So just making sure that some of the institutions that are government funded that touch every body in the community are able to start sharing that knowledge early.  The community gardens too. So community gardens, school gardens, all that growing food. So if you want to even take it a step back (there is a backup plan to continue the initiative).  So I think like a combination of having, you know, health care workers that are licensed to do X, Y and Z, but also people who are non health care workers or who just are involved. And as a community health worker, for example, I guess that is a health care worker, but that focus is more on the community education, um, but folks from the community and trusted leaders  even like you mentioned partnering with jewels. Imagine every time they shop they're able to get you know, some type of free fruit or vegetable with a note attached to adjust with some form of simple education. |
| Capture and share local knowledge | Capture local knowledge from implementation sites on how implementers and clinicians made something work in their setting and then share it with other sites | But in terms of how to operationalize cooking classes, there are some health centers that have kitchens that can be tapped into as local spaces. Chicago Family Health Center, where I used to work on the South Side, had a community kitchen. So to identify spaces that have kitchens that you can then use for the cooking demonstrations. Cooking enough, where you do a demonstration that everyone can sit down and eat together so that you go, you cook and you get a good meal from it.  Feeding America…they've done a huge like change of like, like what can be like donated and all these things because usually, you know, for lack of professionalism, we just give our crap, right? But now there's been a huge change in that and I think it's slowly trickling down. |
| Conduct ongoing training | Plan for and conduct training in the clinical innovation in an ongoing way | But like again, the science of this, I would really, we love to see the science of what's the best approach (HP, FG 1)  I was just laughing. I was like, apparently you need to retrain providers (HP, FG 1) |
| Conduct educational meetings | Hold meetings targeted toward different stakeholder groups (e.g., providers, administrators, other organizational stakeholders, and community, patient/consumer, and family stakeholders) to teach them about the clinical innovation | I mean, there is different programs. One is for any patient we give like force, vegetables and all those but the shop printing, and all these are mainly specifically for what they can eat. And you know, and what do you need to buy when they go to the store? And all those things. So it's really about one on one education. And right, kind of shopping for the food for the phone. Yeah (HP, IDI 2) |
| Revise professional roles | Shift and revise roles among professionals who provide care, and redesign job characteristics | it's really it's just getting to know all the patients and having people be made aware. I try to communicate with the MAs a little more and kind of get, you know, get them on board to kind of help out if we have time, you know, with those kinds of things. I'm really, really impressed with our teams, you know, and I think that they would be up for more of that |
| Create new clinical terms | Change who serves on the clinical team, adding different disciplines and different skills to make it more likely that the clinical innovation is delivered (or is more successfully delivered) | what about getting some student volunteers from social work programs to come help, you know, to volunteer. Like they get something, we get something, they have some accountability, right? I don't know…Or nurses from nursing school.  maybe it is like a worker or somebody like a role that does like, looks for all the resources and then like provides them to, like, I'm just thinking about how to like disseminate that, um, by area or something like that, I don't know. |
| Involve patients/consumers and family members | Engage or include patients/consumers and families in the implementation effort | In the pharmacy I do the same thing too. Like after when I'm counseling patients, I'm constantly giving them handouts and even if they don't read it, I feel better that I gave it to them. And maybe next time when they come in, we can talk about it or something like that. Or sometimes I give them homework because I kind of mess around with the patients, you know, we have like a very free open, you know, we talk when I'm not at work. You know, it's professional, but it's kind of like we joke around and have a good time, too. So I give them assignments like, you know, next time you come in, you better tell me the top five things on there, you better do your homework, you know, stuff like that. And they really get a kick out of it and they do it, you know, they like stuff like that. So. Yeah, hand outs and visual aids are super helpful.  I think if we instilled a value in growing things at home and eating things that you've grown at home and I think the Church can help with that. I think. Local restaurants, of course, schools, but ultimately your family.  And like demonstrating the app, you know, somehow, just kind of getting buy-in. Everybody has smart phones now. |
| 7.2. Intervene with patients/consumers to enhance uptake and adherence | Develop strategies with patients to encourage and problem solve around adherence | I'm not sure that the data is actually there for that, actually, that once people know it, hearing it doesn't help them unless they've already committed to change. So working on motivational interviewing for salt intake may be the place to go.  And I thought about this like just literally meeting them where they're at like what's your ability to go to the grocery store? Are there any grocery stores around you, or do you just have a convenience store kinda like nearby, you know, you went and grabbed a pizza, you know, at this place like literally.  one thing as far as, like, healthy eating habits go…there have also been other studies where there's been cooking classes…they brought people in and they like all cooked together. And actually it was like a communal piece too. So it wasn't just like, Hey, I'm going to teach you how to cook to be healthy, that was part of it. But a lot of it was like, we're all going to get together and this is going to be like, so it was more holistic…So that could be also a way, because I just think like the more hands on, like we're usually like "print out my plate", "print out dash diet", but the more you can like go shopping with someone, cook with them, whatever, like make it fun, make it life giving, I think then people associate that more with like something they want to do and that makes them feel good too.  Many things can be addressed by being in the community rather than the clinic reaching out to, you know. We have a good community outreach program where they where we have events to teach them cooking. We are planning to build a nutrition center attached to this clinic where the community can come and eat. Of course they have to pay the money, but healthy food choices are expensive. And my dietician here she takes the people. She's a diabetic educator. She takes the patients out for shopping in one of the local grocery stores. It's called Market where we have an arrangement with them that they give healthy food for like foods, vegetables, all those healthy foods at current use cost to the patient. So we do such a outrageous. And then also we get donations of healthy green leafy vegetables and fruits and vegetables doing certain fruit store, I mean full store, which we donate to our patients, too. So those, I think, and things can really help for them to be more eat, more healthy food. |
| 7.3. Prepare patients/consumers to be active participants | Prepare patients/consumers to be active in their care, to ask questions, and specifically to inquire about care guidelines, the evidence behind clinical decisions, or about available evidence-supported treatments | And eating habits are formed young. They are really difficult to change when you're older, really difficult. So if you grow up on your healthy snacks, more likely you're going to crave those later. But if you just get diagnosed with X, Y, Z, and then you have to change all your eating habits, that's a big ask. Like that's really, people can do it, but that's a bigger ask. So I totally agree. Get it in the schools, we're already having to provide food for schools, so do it. And I say make it like whatever, like the gardens, all that good stuff. There's so many schools that do that now.  I'm wondering if there's like basic boxes or basic kits of, "here is your kitchen starter kit, here is a mixing spoon, here's a big pot, a little pot or pan". Like, what are the most basic things you need at the most basic prices in order to begin implementing the "this is how you cook at home" advice and then who can we get to fund it and provide instructional booklets and training? Maybe in high schools so that people as they become independent, are able to have that in their tool kit.  The other thing that I'm just thinking culturally is, because it's a question of do you have access to your own kitchen? If you do have a kitchen, do you have the pots and pans you need to cook for yourself? If you do have the pots and pans, you need to cook for yourself, and the oil and the basic ingredients, do you know how to put them together to make food? Has this been taught? Has this been trained? Does it get handed down from parents to children? Has there been that knowledge dropped? And if so, how do you recreate the knowledge of how to cook while having the basic supplies necessary to cook rather than eating the processed foods and the fast foods |
| 7.5. Use mass media | Use media to reach large numbers of people to spread the word about the clinical innovation | And some people don't even realize anything like we know about salt, they don't think about that at all. So I think having like, a whole campaign dedicated to that, um, advertising and all that. |
| 8.1 Fund and contract for the clinical innovation | Governments and other payers of services issue requests for proposals to deliver the innovation, use contracting processes to motivate providers to deliver the clinical innovation, and develop new funding formulas that make it more likely that providers will deliver the innovation | so I think and I know there was initiatives, I don't know if they're still in place to like allow people to use Wick at like farmers markets and like fruits and vegetable gardens and even get like a rebate or get like money back. But I just think more initiatives like that where it's not just like low cost or ideally free, right for these healthier alternatives, but that there also is like an incentive like, oh man, I'm not just going to get this for free, I'm going to get this extra thing. Yeah. So I think just working with, you know, different kind of. But it's hard too, right? For like the food banks and for shelters and things like that because they're not operating on a really big budget. So that's, that's what's tricky. And that's where I think you need like the policy piece to support it. |
| 8.2. Access new funding | Access new or existing money to facilitate the implementation | But again, it needs to be funded, it needs to be seen as health care. It needs to be funded just as much as like big pharma, you know, because it's just as, it's more important but yeah.  Everything that happens needs to be funded through a grant or something because, you know, the clinics are barely trying to keep their doors open. And, you know, they have a lot of patients who are self-pay who really don't get paid for their thing. So, you know, we would have to focus on or you guys would have to focus on finding grants for all of these extra stuff you guys would like to add. |

Note. The category of “Provide iterative assistance” was not coded in the data.

Abbreviations: COMBI-SR. Communication for Behavioral Impact for Sodium Reduction. ERIC. Expert Recommendations for Implementing Change. FG. Focus Group. HP. Healthcare Professional. IDI. In-depth interview. PIR. Potential intervention recipient. RMA. Research Ministry Ambassador.
